# The use of Mendelian randomization to explore the causal consequences of childhood maltreatment: consideration of assumptions and challenges

**DOI:** 10.1101/2025.10.17.25338214

**Authors:** Ka Kei Sum, Amanda M. Hughes, Alexandra Havdahl, George Davey Smith, Laura D. Howe

## Abstract

Mendelian randomization (MR) uses genetic variants as instrumental variables to enhance causal inference. Studies have identified genetic variants related to childhood maltreatment, but interpreting the effects of these variants or assessing the plausibility of MR assumptions is complex. We aim to investigate the feasibility of applying MR to complex social traits using the association between childhood maltreatment and mental health and behavioral outcomes as an example. We explore four potential key concerns: confounding by population phenomena, horizontal and vertical pleiotropy, reverse causality, and selection. For each concern, we demonstrate scenarios where MR studies of childhood maltreatment may be biased using DAGs and critical appraisal of previous MR analyses. For confounding by population phenomena, we further perform within-family genetic analyses in 42,101 parent-offspring trios from the Norwegian Mother, Father and Child Cohort Study (MoBa) to address bias due to family-level processes since childhood maltreatment often occurs within households. Our results showed same-trait shrinkage (11% attenuation of the association between children’s polygenic risk scores of childhood maltreatment (PRS_CM_) and mothers’ report of children’s physical abuse) but not cross-trait shrinkage (children’s PRS_CM_ and children’s mental health and behavioral outcomes) after adjusting for parental PRS_CM_. The lack of cross-trait shrinkage suggests that genetic variants related to childhood maltreatment may be capturing other child-level phenotypes, after adjusting for family-level processes. Mothers’ PRS_CM_ were also associated with mothers’ own maltreatment experiences in childhood and adulthood with similar magnitudes, suggesting these genetic effects are not specific to childhood maltreatment. Due to the complexity involved in the causal chain of childhood maltreatment and it being reported, the interpretation of MR studies for childhood maltreatment is challenging. Other causal approaches should be considered for observational studies of complex social traits.

## Introduction

Childhood maltreatment encompasses physical, emotional or sexual abuse, and neglect. Maltreatment can occur in the home, in an institution, in the community, or, more rarely, by people unknown to the child. It is most commonly perpetrated by parents or caregivers within the household,^1,2^ but in most population-based research, a combined measure of maltreatment is considered, regardless of the perpetrator. Childhood maltreatment is common (global prevalence up to 36%),^2,3^ and is strongly associated with worse physical and mental health outcomes^4^ across the life course. However, given the strong influence of poverty on childhood maltreatment,^5^ it is possible that some of the association between childhood maltreatment and later outcomes reflects confounding by socioeconomic processes. Similarly, parental mental health problems are a risk factor for childhood maltreatment,^6^ and given the known genetic component to mental health, some degree of genetic confounding is possible. Moreover, children with neurodevelopmental conditions such as autism and ADHD may be more likely to experience harsh responses from adults due to evocative gene-environment correlations.^7,8^ Applying causal inference approaches may therefore help us better understand the causal effects of childhood maltreatment itself, over and above the effects of confounding influences. However, causal inference within the field of childhood maltreatment research is challenging.

Mendelian randomization (MR) is an approach that leverages genetic variation to understand phenotypic causation.^9^ It is commonly implemented as an instrumental variable (IV) analysis that uses genetic variants, most commonly single nucleotide polymorphisms (SNPs), as IVs to estimate the causal effect of an exposure on an outcome using observational data.^10^ It has been widely used in the field of epidemiology to improve causal inference because of its potential to overcome one of the major barriers to establishing causality in observational studies, namely unmeasured and residual confounding.^11^ To infer a causal effect of exposure on outcome, three main MR assumptions need to be met.^10^ The assumptions of MR include relevance (genetic IVs are robustly associated with the exposure of interest), independence (there are no common causes between genetic IVs and the outcome of interest), and exclusion restriction (genetic IVs influence the outcome only through the exposure). To quantify the magnitude of a causal estimate, an additional fourth assumption is required.^12–14^ There are two versions of this assumption, each providing slightly different interpretations of causal effects. These include homogeneity (the exposure-outcome association does not depend on the value of the IV) and monotonicity (the direction of IV-exposure association is consistent across all participants). Under the homogeneity assumption, MR estimates the average treatment effect, whereas under the monotonicity assumption (a weaker version of homogeneity), MR estimates the local average treatment effect. Depending on the research question, researchers should adopt the version that is most plausible in their context. MR also makes implicit assumption about consistency, a key criterion in establishing causal inference, which states that the outcome under a given exposure value is the same regardless of how that exposure is achieved.^15^ In MR, this assumption is known as the gene-environment equivalence principle.^14,16,17^ It requires changes in the exposure due to genetic variation to have the same effect on the outcome as changes induced by modifiable or environmental factors.

MR was initially conceived to study exposures with genetic proxies that have a well-defined biological function. However, due to the proliferation of large genome-wide association studies (GWAS), SNPs have been identified relating to a wide variety of phenotypes, and MR is increasingly used for complex traits where the functional relevance of genetic proxies is less well understood. This makes it more likely that MR assumptions are violated. While the relevance assumption can be empirically tested, independence, exclusion restriction, and gene-environment equivalence assumptions are challenging to falsify.^18^

Many^12,13,19,20^ have previously explored scenarios in which genetic instruments are associated with multiple traits in MR and discussed when these scenarios may bias MR results and what are the appropriate causal interpretations. Some of these scenarios include confounding, horizontal and vertical pleiotropy, reverse causation, and selection. Complex social traits are often interlinked and clustered. This means genetic variants associated with one complex social trait are often associated with others. Genetic associations of complex social traits in unrelated individuals such as educational attainment and family socioeconomic position (SEP) have been shown to reflect more than direct genetic effects alone and also include indirect effects due to population phenomena such as population stratification, assortative mating, and dynastic effects.^21–24^ Population stratification happens when the genotype-phenotype associations are confounded by population structure, which is systematic differences in allele frequencies between ancestral subpopulations, often after being segregated by geographical location.^21^ Dynastic effects refer to the indirect influence of parental genotype on offspring phenotype through its expression in the parental phenotype,^21^ sometimes called “the nature of nurture.”^25^ This phenomenon is also a form of gene-environment correlation,^21^ most closely resembling passive gene-environment correlation,^22,26^ which states that the parents’ genotypes shape both the child’s genotype and the environment they provide, thus affecting the child’s phenotype.^27^ Other than parental indirect genetic effects, the term is sometimes used more broadly to reflect the social stratification processes within or across generations, for example, effects from siblings, relatives, or grandparents.^28^ Meanwhile, assortative mating arises when couples choose partners who are more similar to themselves than would be expected by chance with respect to phenotypes such as height and education.^29^ When assortative mating happens related to phenotypes that are genetically influenced, couples are likely to be more genetically similar than is typical across the population.^21^ In the context of MR, these population phenomena can induce spurious correlations through confounding between genotypes and phenotypes, violating the independence assumption, in turn biasing MR estimates.^14,24,28^ This potential bias is illustrated by results of within-family MR studies of complex traits, which by holding parental genotype constant are robust to the influence of population stratification, assortative mating, and dynastic effects. MR studies of unrelated individuals suggest a causal effect of body mass index (BMI) on educational attainment, but within-family MR studies show a null effect, suggesting the association in a standard MR analysis reflects bias from population structure and family-level processes.^28^

MR analyses have previously been used to investigate the relationships between childhood maltreatment and mental health and behavioral outcomes using SNPs identified from a GWAS of childhood maltreatment.^30^ However, SNPs identified for complex social traits such as childhood maltreatment are likely to be correlated with other social, environmental, and mental health traits, making it highly improbable that the independence, exclusion restriction, and gene-environment equivalence assumptions are met. Since childhood maltreatment is something that happens to a child, perpetrated by another person, it is implausible that a child’s genetic variants directly cause maltreatment. Rather, they may influence individual traits (e.g. impulsivity, aggression, emotional regulation) that alter the likelihood of maltreatment occurring through social interactions, or that alter its reporting/recollection. These mechanisms align with evocative gene-environment correlation, where the child’s genotype influences his / her behavior, which evokes reactions from others. Moreover, estimates from 171 countries indicate that the most common perpetrators of childhood maltreatment are household members of both sexes and various ages, except for sexual abuse.^1^ Similarly in high income countries, approximately 80% of maltreatment cases involve a parent or other caregiver, meaning maltreatment can be considered a family-level process for many children.^2^ This suggests MR studies of childhood maltreatment in unrelated individuals may be confounded by parental genotype (*i.e*., dynastic effects), reflecting passive gene-environment correlation.

Hence, we aim to explore the feasibility of applying MR to study complex social traits, using the association between childhood maltreatment and mental health and behavioral outcomes as an example. We explore four potential key concerns: confounding by population structure, parental genotype (*i.e*., dynastic effects, passive gene-environment correlation), and assortative mating; horizontal and vertical pleiotropy; reverse causality; and selection. For each of these concerns we (1) demonstrate scenarios where MR studies of childhood maltreatment and mental health and behavioral outcomes may be biased using directed acyclic graphs (DAGs), and (2) critically appraise previous MR analyses of childhood maltreatment.^30^ For confounding by population structure, parental genotype, and assortative mating, we additionally perform within-family analysis using genotype data from parent-offspring trios in the Norwegian Mother, Father, and Child Cohort Study (MoBa) to explore bias due to family-level processes.

## Key concerns that may bias MR studies of childhood maltreatment

### 1. Confounding by population structure, parental genotype, and assortative mating

Previous literature has demonstrated how population phenomena such as population stratification between ancestral groups, dynastic effects, and assortative mating may bias genotype-phenotype associations^21,24^ and estimates from MR analysis of unrelated individuals.^13,24,28,31^ All these phenomena may bias MR analysis conducted in samples of unrelated individuals by inducing spurious association between the genetic IV and the outcome through confounding.^21^ MR analysis in family-based samples of related individuals – either sibling pairs, or mother-father-child ‘trios’ - is more robust to the impact of these family-level phenomena, because the genotype of other family members can be controlled for.^24,28,31^

Figure 1 demonstrates how population structure, dynastic effects, and assortative mating may bias observed associations in MR studies of childhood maltreatment and mental health and behavioral outcomes.

**Figure 1.**
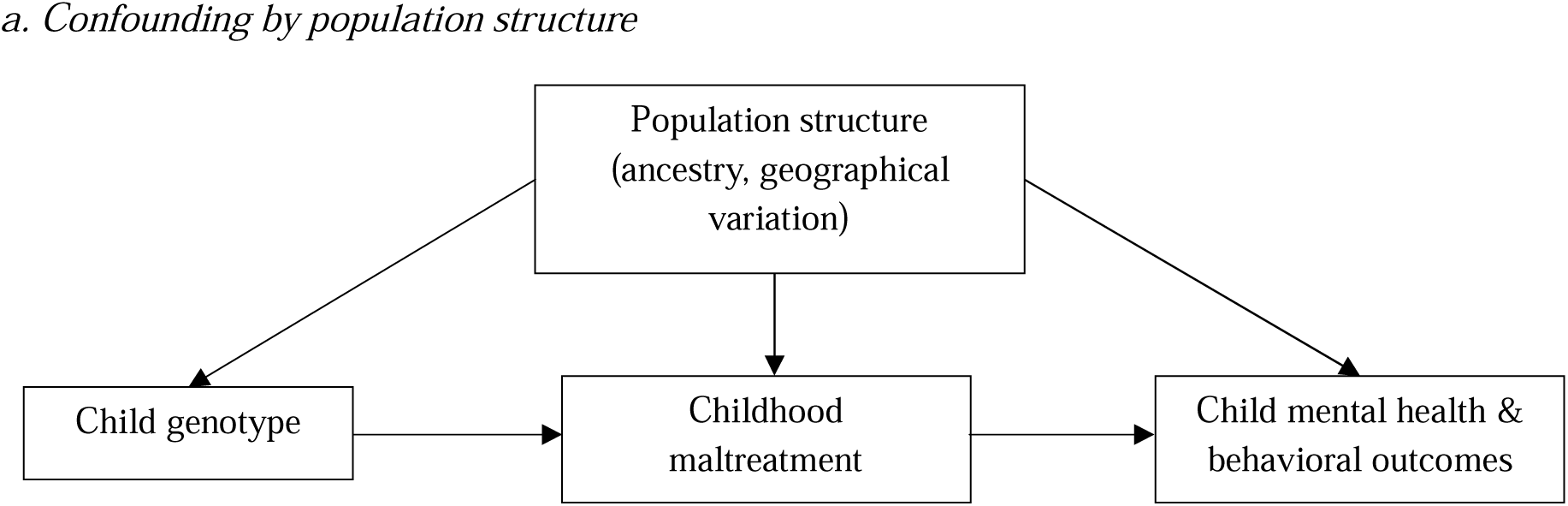
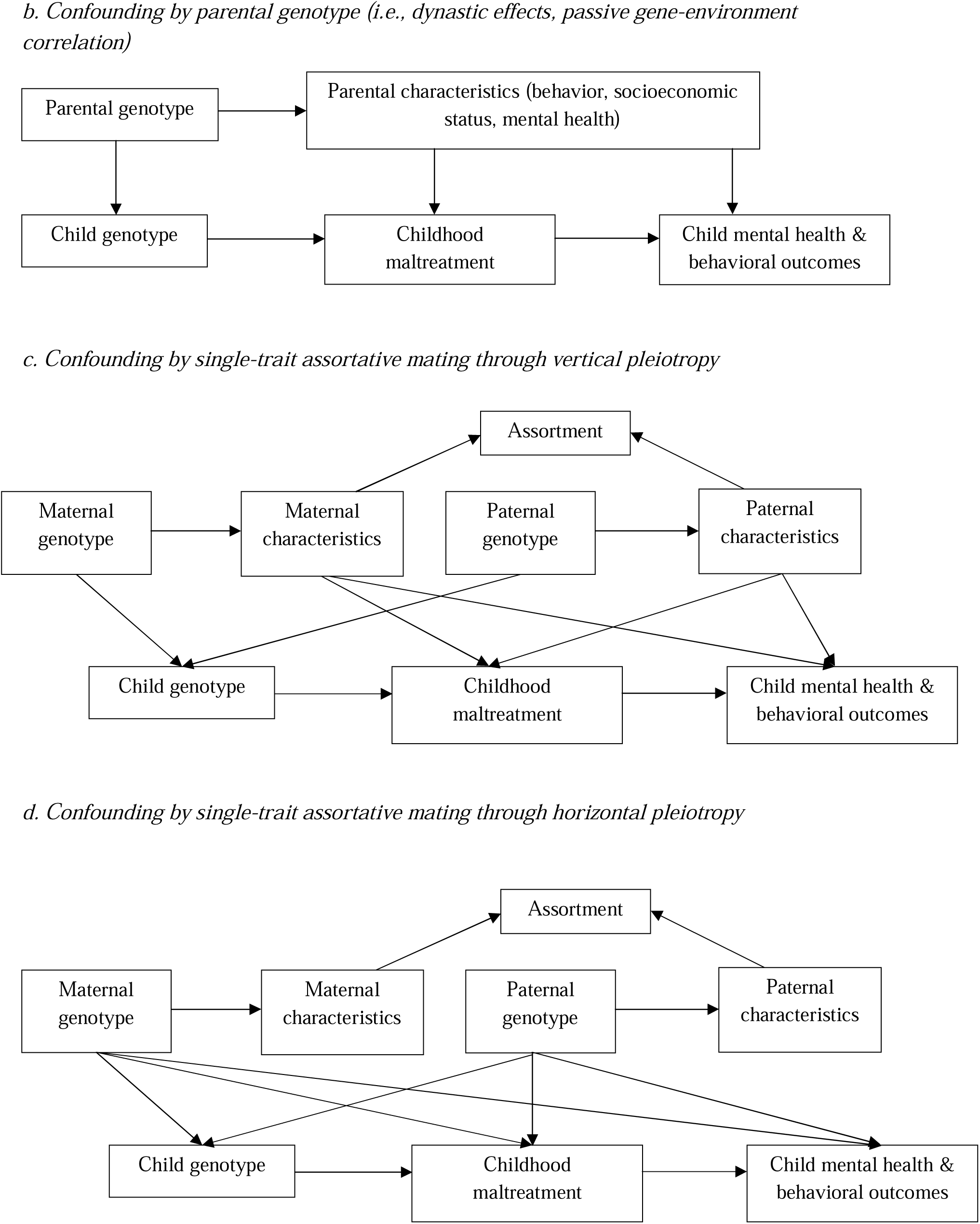
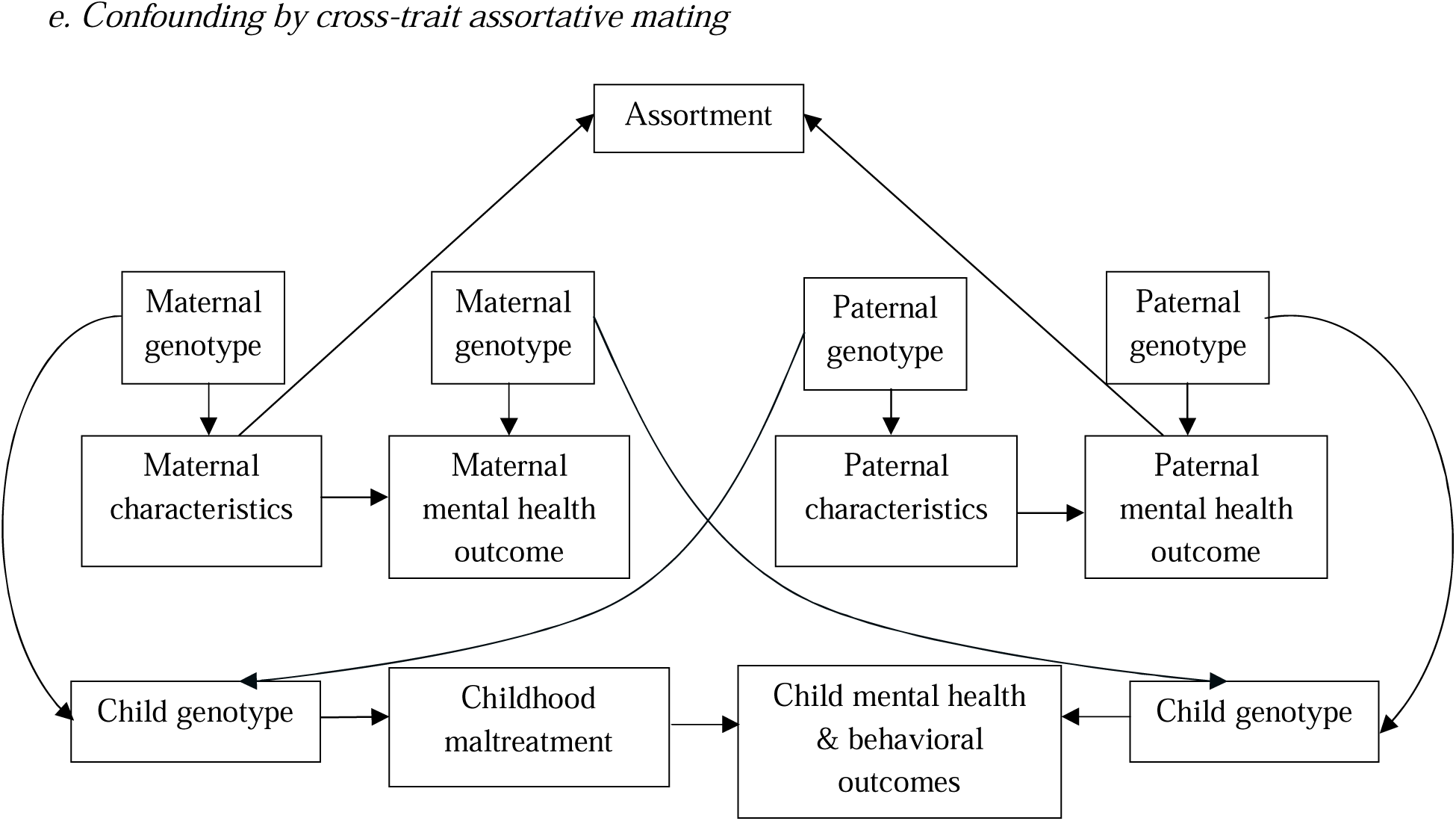
Directed acyclic graph (DAG) representations of confounding by population phenomena. The arrow from child genotype to childhood maltreatment is shown to illustrate potential causal structures, but we do not assume a true direct genetic effect of the child genotype on maltreatment. (a) Child genotype-outcome association is confounded by population structure such as ancestral or geographical variation. (b) Child genotype-outcome association is confounded by parental genotype and parental characteristics / the environment they provide. (c), (d) and (e) Child genotype-outcome association is confounded by single-trait and cross-trait assortative mating. Single-trait assortative mating does not always bias MR estimates. Bias by single-trait assortative mating only happens when the trait under assortment is genetically correlated with both the exposure and outcome, either through (c) vertical or (d) horizontal pleiotropy. All the mechanisms described are not mutually exclusive and multiple mechanisms can happen simultaneously. Unmeasured confounders are not illustrated.

#### 1.1. Previous MR analyses of childhood maltreatment

There have been several MR studies of childhood maltreatment published on physical and mental health outcomes.^30,32–39^ Here, we consider the first of these studies by Warrier et al.^30^ A GWAS meta-analysis of childhood maltreatment (N=185,414) was first performed by the authors using data from five datasets. Variables used to define childhood maltreatment varied across the five datasets and information about perpetrators was usually not specified. They subsequently did a bidirectional MR study investigating the impact of childhood maltreatment on mental health and behavioral outcomes, and vice versa. A ‘two-sample’ MR, it drew on summary statistics from the GWAS of both childhood maltreatment and mental health and behavioral outcomes. Standard inverse-variance weighted (IVW) analysis suggested an impact of childhood maltreatment on depression (beta=0.59, SE=0.15, p=3.63 x 10^-5^) and bidirectional associations of childhood maltreatment with schizophrenia (forward MR of childhood maltreatment on schizophrenia: beta=1.17, SE=0.27, p=1.35 x 10^-5^; reverse MR of schizophrenia on childhood maltreatment: beta=0.05, SE=0.006, p=4.83 x 10^-15^) and ADHD (forward MR of childhood maltreatment on ADHD: beta=1.04, SE=0.36, p=4.02 x 10^-3^; reverse MR of ADHD on childhood maltreatment: beta=0.08, SE=0.02, p=1.22 x 10^-4^).

As summary statistics are usually derived from GWAS in different populations, failure to account for population phenomena in those GWAS will violate the independence assumption and bias MR analyses. While all the GWAS included by the authors for both SNP-exposure and SNP-outcome associations found minimal evidence of genomic inflation due to population structure after considering the intercept from linkage disequilibrium score regression (LDSC), removing genetic outliers, and adjusting for principal components, bias due to dynastic effects and assortative mating may still have been present and can only be accounted for using a within-family design. For this reason, the authors derived polygenic risk scores (PRS) of childhood maltreatment based on the genome-wide significant SNPs from the population-based childhood maltreatment GWAS and performed within-family analyses of the PGS on retrospective, self-reported childhood maltreatment (total scores range from 0-20) using sibling data from the UK Biobank in a linear mixed effects model. The model included two fixed effects, (1) between-family effect (corresponds to the coefficient of the mean family PRS, *i.e*., mean PRS of the siblings) and (2) between-sibling effect (corresponds to the coefficient of the difference in PRS between the individual sibling and the mean family PRS). They identified a between-sibling (*i.e*., within-family) effect (beta=0.053, SE=0.02, p=0.015) of the PRS on retrospective, self-reported childhood maltreatment, interpreted as the direct genetic effect that is free from familial effects which maps onto active/evocative gene-environment correlations.^26^ This between-sibling effect accounted for 58% of the total effect of the PRS (*i.e*., between-family effect) on childhood maltreatment. However, there was little statistical evidence for passive gene-environment correlation quantified by the difference in between-family and between-sibling PRS effects on childhood maltreatment (beta=0.039, SE=0.026, p=0.13; 42% of the total effect). They also found little attenuation in the variance in prospectively reported childhood maltreatment explained by a PRS for retrospectively reported childhood maltreatment in a separate cohort after adjusting for prenatal parental risk factors including smoking, alcohol consumption, depression, and parental maltreatment, which should partly account for potential dynastic effects. Thus, the authors concluded active or evocative gene-environment correlation to be the main mechanism at play, but they could not completely rule out passive gene-environment correlation.

While a sibling-based design accounts for parental indirect effects, between-sibling estimates can still be impacted by indirect genetic effects of siblings.^24,40^ Also, the two analyses only considered confounding by parental genotype in the PRS-exposure (same-trait) association but not the PRS-outcome (cross-trait) association, in this instance, mental health and behavioral outcomes. This means if dynastic processes affect both childhood maltreatment and mental health and behavioral outcomes (DAG 1b), bias due to dynastic effects would not be corrected for in the analyses performed by Warrier et al.^30^ since there are still open paths from parental genotype to the outcomes.^28^ Previous literature has also shown cross-trait shrinkage in within-sibship MR estimates for body mass index (BMI) genetic variants on educational attainment, age at first birth, and cognitive ability when compared to the population estimates even in the absence of same-trait shrinkage in the GWAS estimates of BMI.^24^ Currently, there are no other MR studies on childhood maltreatment that have used study designs to correct for bias due to population phenomena.

#### 1.2. Empirical study using parent-offspring trio genetic data from the Norwegian Mother, Father, and Child Cohort Study (MoBa)

To account for possible family processes influencing childhood maltreatment and mental health and behavioral outcomes, we performed within-family analysis using genetic data on mother-father-offspring trios in the Norwegian Mother, Father, and Child cohort study^41,42^ (MoBa; see detailed methods in supplementary materials; Supp. Fig. 1). Using a trio-based design to correct for indirect genetic effects has been shown to be less biased compared to other within-family designs.^40^ Briefly, we constructed z-standardized childhood maltreatment PRS for mothers (PRS_M_), fathers (PRS_F_), and children (PRS_C_) using 12 independent genome-wide significant SNPs derived from the GWAS of childhood maltreatment performed by Warrier et al.^30^ We then assessed the association between PRS_M_ and the mothers’ reports of their own childhood and adulthood experiences of maltreatment (physical, sexual, emotional, verbal abuse) and their own traits of anxiety, depression, and ADHD. The same regressions were performed for fathers using PRS_F_. Lastly, mothers’ reports of their children’s exposure to physical abuse, and children’s traits of anxiety, depression, and ADHD were regressed on PRS_C_. We then assessed whether associations of PRS_C_ with the children’s mental health and behavioral outcomes were attenuated after adjustment for parental genotype.

We found statistical evidence of associations between PRS_M_ and mothers’ reports of almost all maltreatment experiences in both childhood and adulthood (all RRs between 1.04-1.09 per SD higher of PRS; Supp. Fig. 2), with effect sizes similar for maternal experiences of maltreatment during childhood and adulthood. However, we did not find statistical evidence of an association between PRS_F_ and fathers’ reports of their own sexual (RR=1.14 [95% CI=0.79, 1.63], p=0.49) and physical abuse (RR=1.09 [95% CI=0.96, 1.24], p=0.17) experiences in the past year during mothers’ mid pregnancies. Fathers’ reports of sexual and physical abuse measured during their partners’ pregnancies were rare, only 0.18% and 1.42% respectively, compared to mothers’ reports (17.81% and 13.95%) during the same period. PRS_C_ was also associated with mothers’ reports of their children’s exposure to physical abuse at age 8 years (RR=1.27 [95% CI=1.02, 1.57], p=0.029; Supp. Fig. 3). After adjusting for parental genotype, the association between PRS_C_ and mothers’ reports of their children’s exposure to physical abuse was partly attenuated (RR=1.13 [95% CI=0.84, 1.53], p=0.42; 11% attenuation; Supp. Fig. 3), showing same-trait shrinkage.

Mental health and behavioral outcomes were log-transformed because of their right-skewed residual distributions and were dichotomized if log transformation did not result in normally distributed residuals. In terms of PRS associations with mental health and behavioral outcomes (Supp. Fig. 4), there was statistical evidence of associations between PRS_M_ and mothers’ reports of their own depression and anxiety traits during pregnancy week 30 (RR=1.05 [95%CI=1.00, 1.11], p=0.03), PRS_C_ and mothers’ reports of children’s depression (RR=1.25 [95%CI=1.09, 1.45], p=0.002) and ADHD traits (expected ratio of geometric means=1.02 [95%CI=1.004, 1.03], p=0.01) at age 8 years, and PRS_C_ and children’s reports of their own depression traits (expected ratio of geometric means=1.02 [95%CI=1.003, 1.03], p=0.02) and mothers’ reports of children’s ADHD traits (expected ratio of geometric means=1.04 [95%CI=1.03, 1.06], p=2.38 x 10^-6^) at age 14 years.

Nevertheless, we found little to no attenuation in the estimates between PRS_C_ and children’s mental health and behavioral outcomes after adjusting for parental genotype (*i.e.*, absence of cross-trait shrinkage; Supp. Fig. 5). We also found no strong statistical evidence of associations between PRS_M_ or PRS_F_ and children’s mental health and behavioral outcomes before or after accounting for PRS_C_, suggesting minimal parental indirect effects and that the child genotype does not reflect parental genotype (Supp. Fig. 6). We performed sensitivity analyses by regressing mothers’ childhood maltreatment experiences on PRS_C_ as negative control and found that PRS_C_ did not predict mothers’ childhood maltreatment experiences with or without adjusting for PRS_M_ (Supp. Fig. 7). We also repeated our analyses using a PRS that was constructed with the inclusion of all SNPs from the childhood maltreatment GWAS (as opposed to restricting to genome-wide significant SNPs) and observed similar results (Supp. Fig. 8).

## 2. Pleiotropy

One of the biggest threats to the validity of MR studies is pleiotropy, where genetic variants are associated with multiple phenotypes. There are different types of pleiotropy, mainly categorized into horizontal and vertical pleiotropy. Horizontal pleiotropy happens when there is an alternative pathway from the genetic IV to the outcome that is not through the exposure, thereby violating the exclusion restriction assumption and causing bias in MR estimates. Horizontal pleiotropy is further divided into uncorrelated and correlated pleiotropy. Uncorrelated pleiotropy occurs when genetic variants influence the exposure and the outcome through distinct pathways and the pleiotropic effect on the outcome is independent from the genetic effect on the exposure.^43^ Meanwhile, correlated pleiotropy occurs when genetic variants influence the exposure and the outcome through a confounder such as a shared pathway and the pleiotropic effect is correlated with the effect of the genetic variants on the exposure.^43^ Correlated pleiotropy is a consequence of heritable confounding, because the confounder is driven by the same genetic variants affecting the exposure, and thus “heritable.”^20^ On the other hand, vertical pleiotropy happens when genetic variants are associated with phenotypes that are on the same causal pathway as the exposure and the outcome. It has been shown that vertical pleiotropy does not usually bias MR estimates except when it happens through misspecification of the primary phenotype.^14^ When genetic variants are associated with a phenotype that is upstream of the exposure (*i.e.,* misspecification of the primary phenotype), bias in the MR estimates of the exposure on the outcome happens through the alternative path from the true exposure to the outcome. Misspecification of the primary phenotype is a case of heritable confounding, which leads to correlated pleiotropy.^20^

We present four scenarios of how the genetic IV of childhood maltreatment may influence child mental health and behavioral outcomes via independent pathways and potentially bias MR analysis (Figure 2). In DAGs 2a and 2b, child genotype influences both childhood maltreatment and child mental health and behavioral outcomes independently but in DAG 2b, child genotype also influences child mental health and behavioral outcomes via childhood maltreatment. In DAG 2a, there is no true causal effect of childhood maltreatment on child mental health and behavioral outcomes but they both share the same genetic predictor. DAG 2c and 2d could arise in the situation when a heritable confounder exists between the exposure and the outcome. In DAG 2c, some of the genetic variants in the child genotype are associated with the childhood maltreatment through the confounder, whereas in DAG 2d, all genetic variants in the child genotype are associated with childhood maltreatment through the confounder. DAG 2d demonstrates a scenario that childhood maltreatment is a proxy for a separate underlying causal trait that affects mental health and behavioral outcomes, for instance relating to the child’s own characteristics that predispose the child to mental health conditions. When this underlying causal trait is related to child behavior or personality, this scenario represents evocative gene-environment correlation, where child genotype influences child behavior or personality which in turns influences the child’s risk of being maltreated, thus affecting their risk of developing mental health and behavioral outcomes. For example, children with ADHD and autism may be more likely to experience harsh parenting because of behaviors perceived as challenging.^7,8^ The evocative responses may even be mediated by an active gene-environment correlation, where the genetically influenced child characteristics shape the family environment that affects their risk of maltreatment. It is also possible that the genetic variants may be related to the child’s characteristics associated with differential reporting of childhood maltreatment instead of childhood maltreatment itself. Even if maltreatment may not be self-reported, parental or caregiver response may differ based on what the child has disclosed to them. In DAG 2d, the testing of the null hypothesis of no causal relationship between the exposure and the outcome remains valid because some aspects of the underlying causal trait are being captured by the mis-specified primary phenotype. However, the causal effect estimate will be biased.^12^

**Figure 2.**
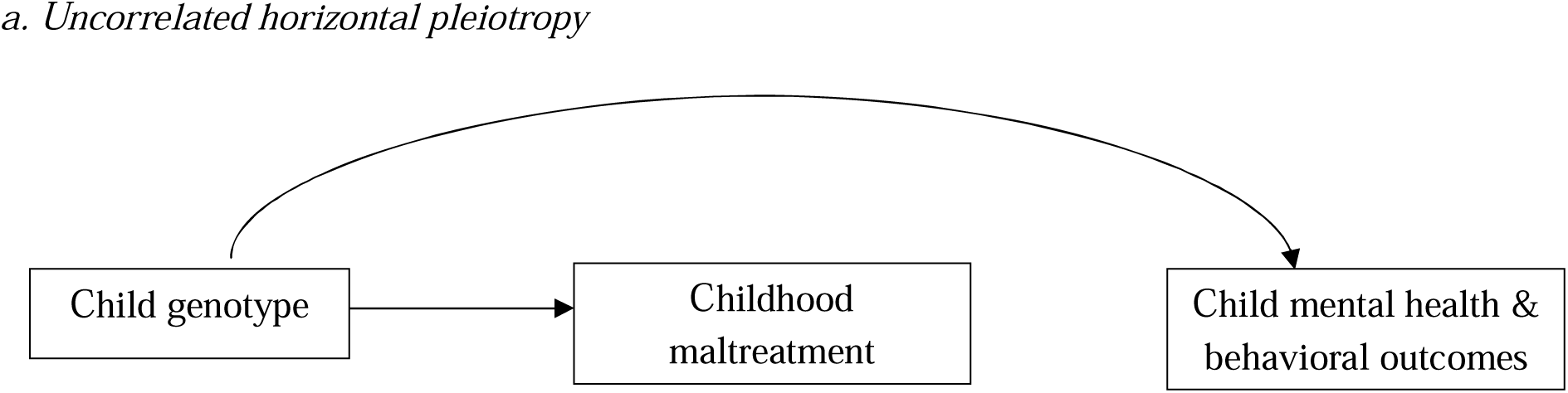
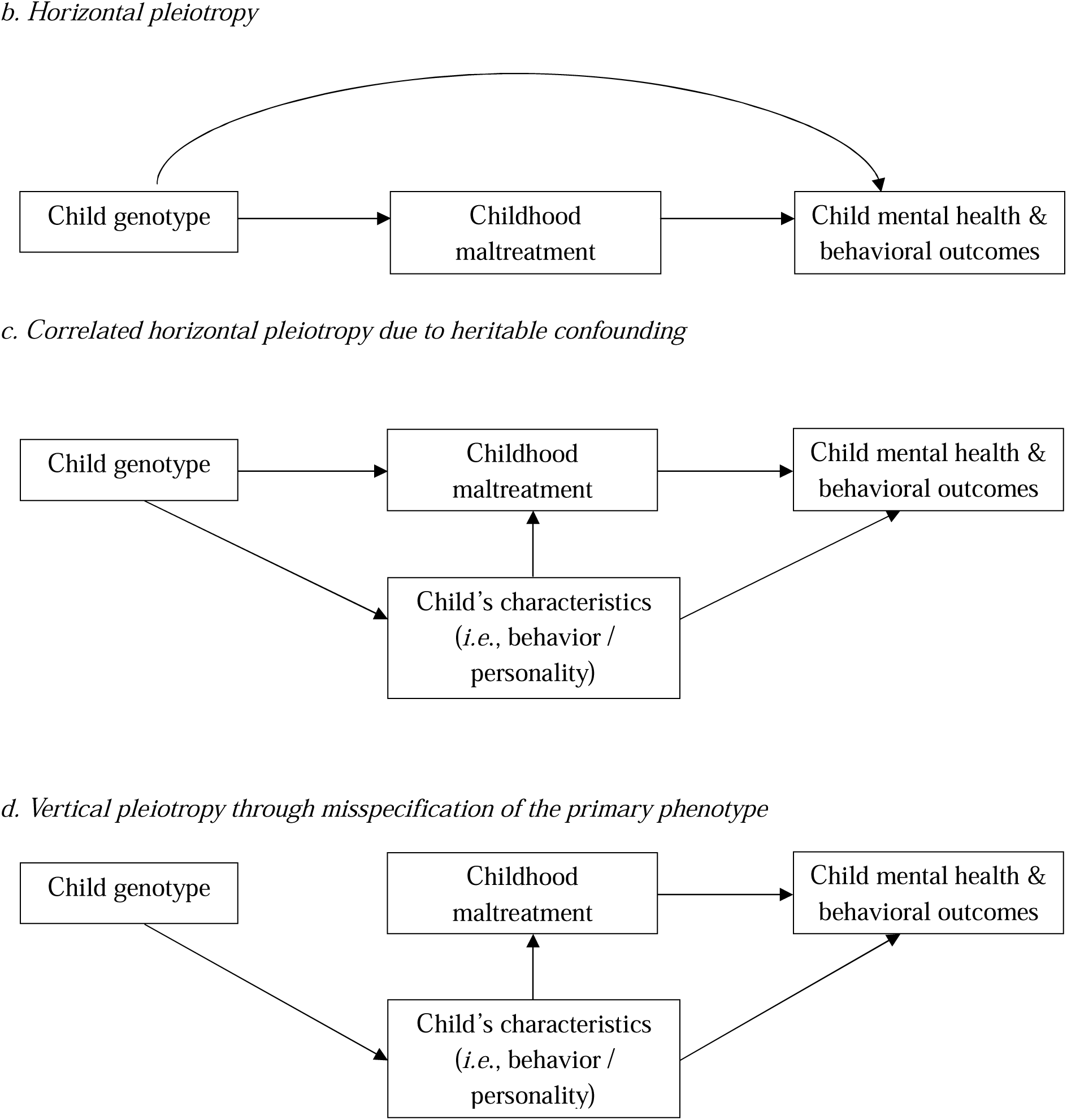
Directed acyclic graph (DAG) representations of non-exhaustive scenarios of horizontal pleiotropy. The arrow from child genotype to childhood maltreatment is shown to illustrate potential causal structures, but we do not assume a true direct genetic effect of the child genotype on maltreatment. (a) Horizontal pleiotropy happens when genetic variants believed to influence childhood maltreatment are also associated with child mental and behavioral outcomes, but there is no causal relationship between childhood maltreatment and the outcomes of interest. This represents uncorrelated pleiotropy, where the effects of genetic variants on the exposure and the outcome are independent. (b) Horizontal pleiotropy happens through the same mechanism as (a) but there is causal relationship between childhood maltreatment and child mental health and behavioral outcomes. This can be uncorrelated or correlated depending on whether the genotype-outcome path is independent of or has shared mechanism with the genotype-exposure path. (c) Genetic variants believed to influence childhood maltreatment are associated with a confounder (i.e., child’s behavior / personality). This represents correlated horizontal pleiotropy, where the pleiotropic effect is correlated with the genetic variants’ association with the exposure. It is a case of heritable confounding because the confounder is a shared heritable factor that is genetically driven. (d) This is essentially (c) but without the arrow from child genotype to childhood maltreatment. Here, the genetic variants are primarily associated with the confounder but not with childhood maltreatment. This represents a subtype of vertical pleiotropy, which biases MR estimates. Misspecification of primary phenotype is also a case of heritable confounding. Unmeasured confounders are not illustrated.

**Figure 3.**
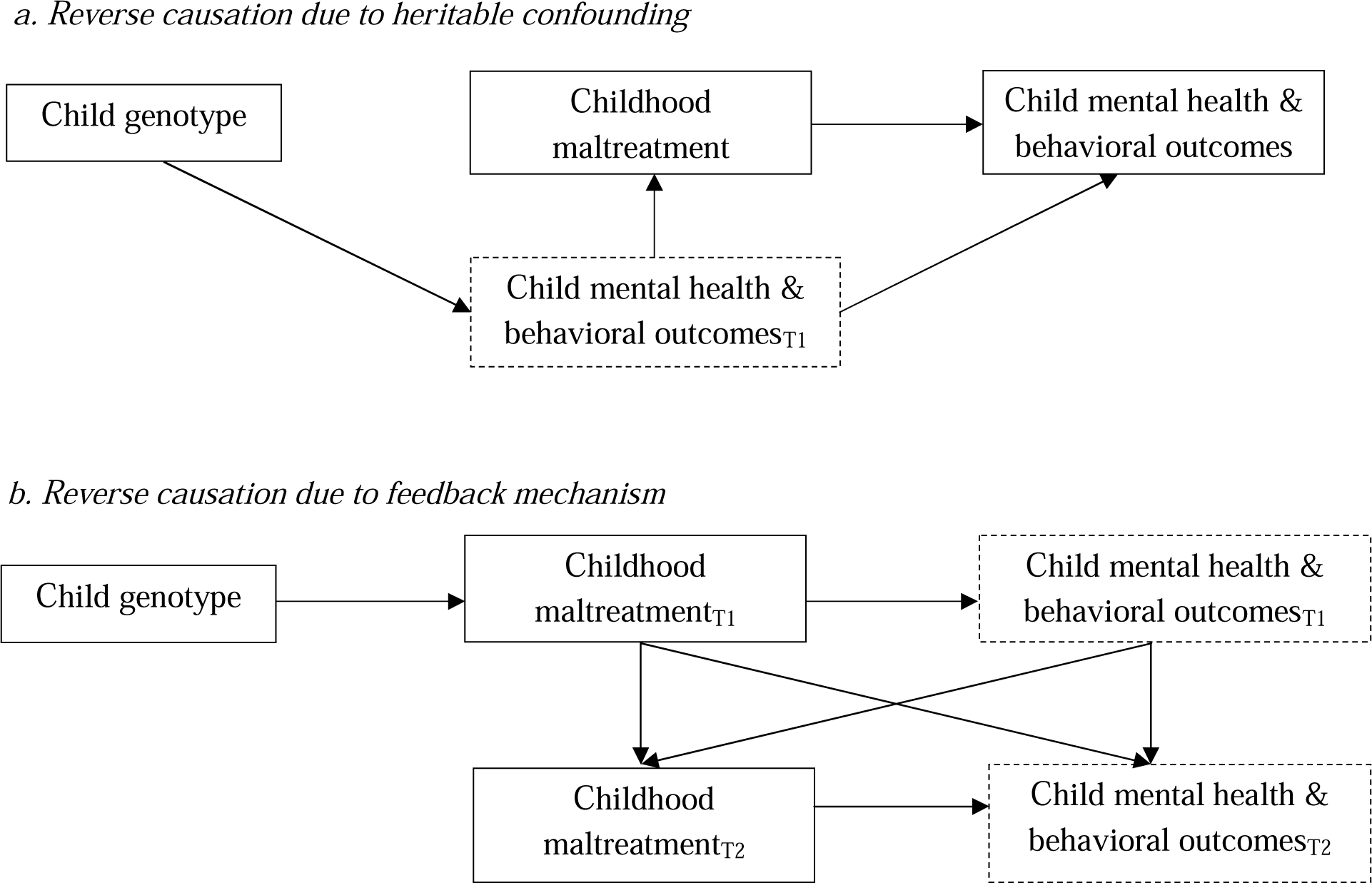
Directed acyclic graph (DAG) representations of reverse causation. The subscript T refers to timepoint. The arrow from child genotype to childhood maltreatment is shown to illustrate potential causal structures, but we do not assume a true direct genetic effect of the child genotype on maltreatment. (a) Reverse causation happens due to genetic variants primarily affect child mental health and behavioral outcomes rather than childhood maltreatment. This is essentially the structure of heritable confounding as presented in DAG 2d. The heritable confounder here is the outcome itself. However, it can also be some psychopathological processes involved before the outcome is developed (dashed box). (b) Reverse causation occurs as a result of feedback mechanism, where there is a bidirectional relationship between childhood maltreatment and child mental health and behavioral outcomes. The variables in the dashed box can be replaced by other psychopathological processes involved before the outcome is developed. Unmeasured confounders are not illustrated.

**Figure 4.**
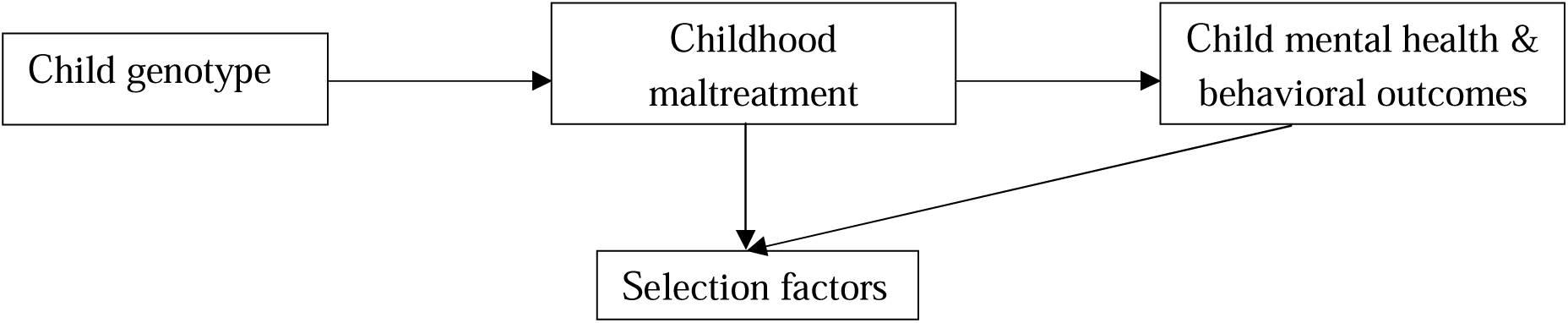
Directed acyclic graph (DAG) representations of selection. The arrow from child genotype to childhood maltreatment is shown to illustrate potential causal structures, but we do not assume a true direct genetic effect of the child genotype on maltreatment. Both childhood maltreatment and child mental health and behavioral outcomes influence the child’s selection into the study. Conditioning on the selection factors can cause spurious association between the child genotype and the outcome even if there is no true causal effect. Unmeasured confounders are not illustrated.

### 2.1. Previous MR analyses of childhood maltreatment

Performing multiple pleiotropy-robust MR methods that have different assumptions has been recommended to increase the credibility of causal claims.^44^ Weighted median MR relaxes the IV assumption by allowing less than half of the variants to be invalid instruments. MR-Egger allows all the genetic instruments to have pleiotropic effects and includes an intercept term to estimate the average pleiotropic effect. However, it relies on the Instrument Strength Independent of Direct Effects (InSIDE) assumption, where variant-exposure associations should not be associated with the pleiotropic paths, which is often not plausible. Meanwhile, MR-PRESSO is a variation of inverse-variance weighted (IVW) regression which involves sequential removal of variants that may have substantially different ratio estimates (*i.e.*, potential pleiotropic variants) from the rest of the variants. Weighted median MR and MR-PRESSO are robust to outliers but not MR-Egger. While MR-PRESSO is efficient with valid IVs, it is likely to have high false positive rate in the presence of invalid IVs. Another important caveat to all the pleiotropy robust methods is that they can only account for uncorrelated pleiotropy but less so for correlated pleiotropy.^20,43^

Warrier et al.^30^ replicated the results of their standard IVW analysis using both median weighted MR and MR-PRESSO, whereas MR-Egger only showed statistical evidence for bidirectional effect for schizophrenia. Evidence of associations between childhood maltreatment and major depressive disorder and ADHD were not replicated in MR-Egger. Statistical power in MR-Egger analyses is generally lower than for other MR methods, which could explain these discrepancies.

Furthermore, their leave-one-out analysis (repeating the MR analysis omitting each SNP in turn) did not suggest a particular genetic variant was driving the result. The authors also attempted to understand if childhood maltreatment SNPs might be related to other phenotypes by identifying previous GWAS associations of those SNPs and performing genetic correlations between childhood maltreatment SNPs and other health related phenotypes. They found childhood maltreatment SNPs were previously associated with mental health and neurodevelopment (depression, ADHD, autism, schizophrenia), personality (neuroticism), risk-taking behavior, smoking and cannabis use, cognitive performance, and educational attainment. In genetic correlation analyses, they also found positive genetic correlations with mental health and behavioral outcomes and negative genetic correlations with intelligence and educational attainment. The largest magnitudes of genetic correlations were those with mental health and neurodevelopmental phenotypes including depressive symptoms (r=0.65), ADHD (r=0.56), and autism (r=0.41).

The authors also performed polygenic transmission disequilibrium tests (pTDT),^45^ a within-family PRS-based method that assesses the likelihood of allele transmission of a certain trait from unaffected parents to their affected offspring, by quantifying how much the offspring PRS deviates from the parentally derived expected value. They hypothesized that siblings who are discordant for neurodevelopmental conditions such as autism or ADHD may prompt differential parental responses, thus indicating evocative gene-environment correlation. They found over-transmission of the childhood maltreatment PRS (*i.e*., where offspring inherited a higher value of the childhood maltreatment PRS than expected by chance) in autistic children in two cohorts, suggesting the presence of evocative gene-environment correlation. Yet, there was no statistical evidence of a relationship between childhood maltreatment and autism in their MR results. The authors suggested this might have been due to weak instrument bias and underlying heterogeneity in autism. They also mentioned the observed over-transmission could arise in the event where childhood maltreatment and autism were driven by a common cause even in the absence of a true causal relationship between the two.

Despite Warrier et al.’s best effort to comprehensively consider potential instrument validity and pleiotropic effects by performing pleiotropy-robust MR methods and additional sensitivity analyses, it is challenging to completely tease apart the scenarios depicted by the DAGs presented in Figure 2 since they may be statistically indistinguishable. In addition, a recent simulation study^20^ has shown that all standard pleiotropy-robust methods will be biased in the presence of correlated pleiotropy caused by heritable confounding, especially if a large proportion of the genetic variants for an exposure are associated with a confounder of the exposure and the outcome. Given the large magnitudes of genetic correlations observed between childhood maltreatment and mental health and behavioral outcomes,^46,47^ it is also possible that childhood maltreatment may have shared etiology with these outcomes, even in the absence of a true causal effect.^12,44^

## 3. Reverse causation

Reverse causation can primarily happen under two scenarios, (1) when the genetic instruments used for the exposure primarily influences the outcome, which in turn influences the exposure of interest (DAG 3a) and (2) due to a feedback mechanism, where the exposure affects the outcome and the outcome affects the exposure at a later time point (DAG 3b).^48^DAG 3a is essentially the same representation of DAG 2d but here the heritable confounder can be the outcome itself or the psychopathological processes preceding the outcome. For example, some of the psychopathological processes that affect individuals who experience childhood adversity are emotional dysregulation and cognitive impairment.^49^ These processes are known to be central to self-regulation and decision making. As discussed in section 2.1, childhood maltreatment SNPs were highly correlated with mental health and behavioral outcomes and processes involved in psychopathology. Children who have impairment in these processes to begin with may influence their risk of being maltreated and developing mental health and behavioral outcomes. Under the feedback mechanism scenario, it is also possible that there exists a bidirectional relationship between childhood maltreatment and these psychopathological processes. Children who are experienced maltreatment may experience impairment in emotional and cognitive processing, perpetuating their vulnerabilities to be maltreated. Bidirectional relationships between childhood maltreatment and mental health and behavioral consequences can be explained by passive or evocative gene-environment correlations. Under both scenarios, since the outcome or development process influences the exposure, genetic variants for the outcome or development process are also associated with the exposure. In large enough sample sizes, genetic variants associated with the outcome or development process will be detected as significantly associated with the exposure in a GWAS of the exposure. In such cases of reverse causation, MR effect estimates are expected to be biased.

### 3.1. Previous MR analyses of childhood maltreatment

Bidirectional MR can be used to orient the causal direction of effect between two traits using two corresponding genetic instruments. To correctly identify causal directionality, the two genetic instruments should be independent from each other. This is as indicated by a Steiger directionality test, which compares the proportion of variance or pseudo-variance (for categorical factors) in the exposure and outcome and removes genetic variants that have greater association with the outcome compared to the exposure. Warrier et al.^30^ performed bidirectional MR to assess the effect of childhood maltreatment on mental health and behavioral outcomes and vice versa. Steiger directionality tests were also performed by the authors in their bidirectional MR, supporting the causal direction indicated by all MR analyses. However, results from a Steiger test may be erroneous in the presence of differential measurement error, unmeasured confounding, and large differences in sample sizes between the exposure and outcome GWAS^50,51^ and detection of the primary influence of genetic variants is not guaranteed.^48^ In most cases, Steiger test is over-permissive of the genetic variants included because it is generally implausible for a genetic variant to statistically explain more than 10% of the variance in the outcome compared with the exposure. For example, even for a strong risk factor such as smoking accounts for only around 10% of the pseudo-variance in lung cancer mortality in a cohort.^52^ Sensitivity analyses to assess the impact of a range of samples sizes, varying levels of measurement error, and unmeasured confounding can be carried out to further understand these sources of bias.^50,51^ Given the challenges of differential measurement error and unmeasured confounding, even in the event where bidirectional MR and Steiger directionality tests may detect some associations in both directions, it is also possible that the observed effects may be indicating shared etiological pathways in the absence of a true causal relationship, *i.e*., heritable confounding. Besides the MR analysis done by Warrier et al., only two^35,37^ of the other eight MR studies on childhood maltreatment performed Steiger tests. However, none performed the sensitivity analyses suggested above to assess the potential biases that may invalidate Steiger test.

## 4. Selection

Conditioning on common effects (or descendants) of both exposure and outcome can create spurious exposure-outcome associations, which is also known as collider bias.^53^ Selection bias is a specific case of collider bias when the characteristics of participants selected into the study are colliders, *i.e.* are caused by both the exposure of interest (here, childhood maltreatment) and the outcome of interest (here, child mental health and behavioral outcomes).^53^ Selection bias is common in most epidemiological studies due to challenges in recruiting participants who are representative of the target population, loss to follow-up, and missing information. Moreover, it is especially problematic in biobanks since participants who volunteered biospecimens are often a subset of people who are fundamentally different from the general population.^54^ As such, analyses that involve genetic data are likely to suffer from selection bias. In the case of MR, selection bias invalidates the independence assumption, where the genetic IV and confounders become conditionally dependent, thus biasing MR results by inducing a spurious association between the genetic IV and outcome even in the absence of a true causal effect between exposure and outcome. Selection bias has been shown to bias MR estimates especially for socio-behavioral traits such as education, diet, smoking, and BMI^55^ since these traits are also more likely to be associated with the broader socio-economic factors, which influence selection. The magnitude of bias is also shown to be more severe in the presence of strong selection effects, *i.e.*, strong exposure-selection, exposure-confounder, confounder-outcome, and confounder-selection relationships but less so when selection effects are moderate.^56^

People with complete data on childhood maltreatment and mental health and behavioral outcomes have been shown to have higher socioeconomic position (more educational qualifications, higher household income, less material deprivation) in multiple cohorts,^57–59^ including those included in Warrier et al.^30^ analysis and our empirical analysis in MoBa. Given the strong effects of poverty on the risk of childhood adversity,^60^ selection due to social patterning will likely underestimate the MR effect sizes of childhood maltreatment and mental health and behavioral outcomes. However, despite the likely underestimation of estimates, the directionality of estimates is likely to be consistent as previously shown.^55^ Hence, we expect the true association estimates between childhood maltreatment and mental health and behavioral outcomes to be in the same direction as estimates affected by selection, but with greater effect sizes.

### Potential interpretations and implications of MR studies of childhood maltreatment

Mendelian randomization is a useful tool to uncover causal relationship between two variables using genetic variants when the IV assumptions are adequately met. In most cases, these assumptions are more likely to be met when a trait is primarily influenced by intrinsic biological factors. We attempted to describe scenarios in which MR estimates might be biased and invalid in the context of a complex social trait, exemplified by childhood maltreatment. We demonstrated strategies to overcome potential biases by critically appraising previous MR studies of childhood maltreatment and conducting an empirical study using trio-based data from a large-scale birth cohort in Norway of childhood maltreatment and its association with mental health and behavioral outcomes. Our within-family analyses showed same-trait shrinkage (PRS_C_ and child physical abuse) but not cross-trait shrinkage (PRS_C_ and child mental health and behavioral outcomes) after accounting for dynastic effects by adjusting for parental PRS. This means that dynastic effects (*i.e*., passive gene-environment correlation) partially explained PRS-childhood maltreatment association but not PRS-child mental health and behavioral outcome associations). Contrary to our results, Warrier et al.^30^ found no strong statistical evidence of passive gene-environment correlation in their between-sibling study for the PRS-childhood maltreatment association but only statistical evidence for active / evocative gene-environment correlation. The lack of cross-trait shrinkage in our empirical analysis suggests that genetic variants related to childhood maltreatment may be capturing other child-level phenotypes, after adjusting for family-level processes. Mothers’ PRS were also associated similarly with mothers’ own maltreatment experiences in both childhood and adulthood, suggesting these genetic effects are not specific to childhood maltreatment, consistent with previous finding.^46^ Multiple MR pleiotropy-robust methods and directionality tests led the authors of the original MR study to conclude potential causal effects of childhood maltreatment on mental health and behavioral traits including depression, ADHD, and schizophrenia. However, we argue that the interpretation of such MR findings is extremely challenging due to potential pleiotropic pathways, reverse causality, and selection even in the context of within-family studies that are more robust to population confounding (one of the biggest threats to genetic studies of social traits).

As previously mentioned, childhood maltreatment is a complex social phenomenon that involves multiple people and forces spanning multiple socio-ecological levels. As with most complex social traits including educational attainment, some genetic variants are likely associated with childhood maltreatment because of their associations with other individual and societal factors along the causal chain.^61^ Our analysis only provided snapshots of potential pleiotropic pathways (Figure 2a-2d) that are parts of a bigger social ecosystem. The complexity involved in the development of a social trait has two implications for MR studies.

First, the interpretation of MR findings becomes extremely difficult. In the absence of comprehensive biological knowledge about the genetic variants, positive MR evidence may reflect a combination of causal and non-causal pathways. In particular, the assumption of gene-environment equivalence is unlikely to hold. It is implausible that genetic variants exert a direct effect on childhood maltreatment. Instead, they may operate indirectly through behavioral tendencies or social dynamics that affect how maltreatment occurs or is reported. Consequently, the pathways through which genetic liability gives rise to maltreatment differ from those through which maltreatment arises due to social, economic, or contextual factors. The genetically influenced form of maltreatment may thus differ qualitatively in type, timing, or severity, violating the assumption that the exposure induced by genetic variation is equivalent to that experienced in the broader population. Even if genetic liability increases the risk of maltreatment in one context (e.g., high-stress households), it may be less likely to do so in another (e.g., supportive environments), meaning gene-environment mapping can be highly context dependent. This pattern mirrors the findings for *ALDH2* variants in women, where the expected genotype-exposure association was effectively “zero” where cultural factors result in low alcohol consumption in this group.^62^ Moreover, gene-environment correlations further confound the interpretation of causal estimates. Together, these issues make it difficult to satisfy gene-environment equivalence in MR studies of socially mediated exposures like childhood maltreatment.

Second, the IV assumption of exclusion restriction may rarely or never be met. Since most childhood maltreatment cases are likely to be to be influenced by some upstream processes, heritable confounding is unavoidable. Despite the availability of MR pleiotropy-robust methods to assess instrument validity, many of them require additional assumptions that are untestable and often implausible such as the InSIDE assumption. This means evidence even from MR pleiotropy-robust studies is undermined if additional sensitivity analyses^63^ such as a heterogeneity test, leave-one-out analyses, use of positive and negative control outcomes, colocalization, subgroup analyses, associations with potentially pleiotropic variables, correcting for selection bias, and investigating reverse causation are not performed. Certainly, as described previously, in the presence of heritable confounding that leads to correlated pleiotropy, all standard pleiotropy-robust methods will be biased if a large proportion of the genetic variants are associated with the heritable confounder.^20^ The guidelines developed to perform^63^ and report^16^ MR studies can be useful to help researchers consider whether the research question at hand can be appropriately addressed using MR by thinking through the plausibility of assumptions and challenges in interpretations.

Moreover, the ethical implications of conducting MR studies of childhood maltreatment can be far-reaching. Debates surrounding the risks and benefits of the wider research of social and behavioral genomics (SBG) have been ongoing since such studies began. Recently, The Hastings Center^61^ further classifies certain SBG studies (largely focused on GWAS and PRS) to be of heightened concern especially those that involve sensitive phenotypes, *i.e*., “phenotypes influencing one’s social standing, historically have been part of harmful stereotypes about minoritized groups and threaten to reify the biologization of social identities, or central to a minoritized group’s identity.” They recommended that such studies should be conducted and communicated responsibly to reduce potential harms. We argue that genetic research specifically surrounding GWAS^30,64^ and creation of PRS of childhood maltreatment should fall under the category of sensitive phenotypes since childhood maltreatment is not only highly damaging but can lead to social stigma for individuals who have experienced it. Even in the context of well-conducted and carefully communicated studies, there is a risk that studying the individual-level genetic correlates of childhood maltreatment could add to stigmatization, discrimination, and fatalism at the individual, group, and policy levels.^61^

That said, while using MR to instrument complex social exposures may not be the best approach to give us causal answers for the reasons stated above, MR can sometimes be valuable to assess the extent of confounding. In particular, MR can be used to instrument potential confounders instead of the social exposure itself. An example is the application of MR to understand the determinants of selection into shift work, where genetic instruments for cardiometabolic risk factors are used to estimate the selection effect of these risk factors on shift work.^65^ These estimates are useful to inform the extent to which these risk factors may confound the association between shift work and adverse health outcomes. A second example of using germline genetics to understand confounding relates to interleukin 6 (IL-6) activity effects on high-density lipoprotein (HDL) cholesterol, and how these confound the associations of HDL cholesterol with various health outcomes.^66^ Other genetic methods may be useful to help disentangle causal effects of childhood maltreatment from confounding.

These include family-based designs such as adoption, children of twin, and sibling comparison studies. Some of these designs can also help us to better understand mechanisms of transmission and gene-environment correlation without the use of GWAS and PRS.^67^ However, PRS can also be useful in cases when it is used in a within family design such as the pTDT approach mentioned earlier or sensitivity analyses to account for genetic confounding.^68^ Other non-genetic designs such as marginal structural models and structural nested models that allow modelling of time-varying exposures and confounders are useful for increasing evidence credibility.^69^ Natural experiments that examine the effects of childhood maltreatment through wider social or political events can also help overcome biases and limitations of traditional observational studies.^70^ This is exemplified by the English and Romanian Adoptees study^71^ that examines the long term mental health outcomes of institutional neglect experienced by young children as a result of extreme early life deprivation that occurred during the CeaulJescu regime in Romania before being adopted by UK families.

### Conclusion

The explosion of GWAS has enabled us to study the genetic predisposition of many complex traits and use genetic variants as instruments for MR studies. MR studies are only valid when the instrumental variable assumptions are met, and they are less likely to be met for traits that are extrinsic to an individual, complex, and socially or environmentally driven. In the context of childhood maltreatment, the underlying causal mechanism linking a person’s genotype to their risk of exposure is likely to be complex and multi-factorial. As demonstrated in our study, even in a well-executed MR analysis, the interpretation is extremely challenging due to potential pleiotropic pathways and gene-environment correlations. Established MR pleiotropy-robust methods and sensitivity analyses have their own limitations as certain assumptions remain untestable and often implausible for complex social traits. There are also important ethical considerations regarding GWAS and PRS research on traits which are more common in vulnerable groups or may be subject to stigmatization. We urge researchers to be mindful when implementing MR for complex social traits such as childhood maltreatment and consider other approaches that may help strengthen causal inference in observational studies.

## Acknowledgements

This study includes data from the Norwegian Mother, Father and Child Cohort Study (MoBa) conducted by the Norwegian Institute of Public Health. We are grateful to all the participating families in Norway who take part in this on-going cohort study. For generating high-quality genomic data, we thank the Norwegian Institute of Public Health (NIPH), the HARVEST collaboration, the NORMENT Centre at the University of Oslo, the Center for Diabetes Research at the University of Bergen, deCODE Genetics, the Research Council of Norway, the South-Eastern and Western Norway Regional Health Authorities, the ERC AdG, Stiftelsen KG Jebsen, the Trond Mohn Foundation, and the Novo Nordisk Foundation. This work was performed on the Tjeneste for Sensitive Data (TSD) facilities, owned by the University of Oslo, operated and developed by the TSD service group at the University of Oslo, IT-Department (USIT), using resources provided by Sigma2—the National Infrastructure for High Performance Computing and Data Storage in Norway (UNINETT).

## Data availability

Data from the Norwegian Mother, Father and Child Cohort Study is managed by the Norwegian Institute of Public Health. Access requires approval from the Regional Committees for Medical and Health Research Ethics (REC), compliance with GDPR, and data owner approval. Participant consent does not allow individual-level data storage in repositories or journals. Researchers seeking access for replication must apply via www.helsedata.no.

