## Supplemental materials for "The use of Mendelian randomization to explore the causal consequences of childhood maltreatment: consideration of assumptions and challenges"

**Supplementary materials**

**Content**

1. Within-family genetic analyses	2-3
   a. Study participants	2
   b. Biological samples	2
   c. Genotyping and quality control	2
   d. Maltreatment history	2
   e. Mental health and behavioral outcomes	2-3
   f. Statistical analyses	3
2. Supplementary tables	4-5
   a. Supplementary Table 1 – Maltreatment questions	4
   b. Supplementary Table 2 – Mental health and behavioral traits measurement	5
3. Supplementary figures	6-14
   a. Supplementary Figure 1 – Cohort workflow	6
   b. Supplementary Figure 2 – PRS and maltreatment associations	7
   c. Supplementary Figure 3 – PRS_C_ and childhood maltreatment at Y8 associations adjusting for parental PRS	8
   d. Supplementary Figure 4 – PRS and mental health and behavioral outcomes associations	9
   e. Supplementary Figure 5 – PRS_C_ and child mental health and behavioral outcomes associations adjusting for parental PRS	10
   f. Supplementary Figure 6 – Parental PRS and child mental health and behavioral outcomes associations	11
   g. Supplementary Figure 7 – Negative control: PRS_C_ and mother’s childhood maltreatment history and mental health outcomes	12
   h. Supplementary Figure 8 – Sensitivity analyses	13
   i. Supplementary Figure 9 – Non-genetic associations	14
4. References	15

**Within-family genetic analyses**

***Study participants***

The Norwegian Mother, Father, and Child Cohort Study (MoBa)^1–3^ is a population-based pregnancy cohort that recruited pregnant women living in Norway and attending routine ultrasound examination during week 17 or 18 week of their pregnancies between 1999 to 2008. Women in 112,903 pregnancies were recruited (41% participation rate), which resulted in 114,500 children, 95,200 mothers, and 75,200 fathers in the cohort. For this analysis, we included 42,101 complete parent-offspring trios (42,101 children, 36,618 mothers, and 36,612 fathers; Supp. Fig. 1) with genotype data that passed post-imputation quality control (QC) and available consents to date. The current study is based on version 12 of the quality-assured data files released for research in 2023. The establishment of MoBa and initial data collection was based on a license from the Norwegian Data Protection Agency and approval from The Regional Committees for Medical and Health Research Ethics (REK). The MoBa cohort is currently regulated by the Norwegian Health Registry Act. The current study was approved by The Regional Committees for Medical and Health Research Ethics (14140; 2016/1702).

***Biological samples***

Blood samples were collected from the mothers and fathers at the ultrasound appointments. The mothers had blood drawn again soon after delivery. Umbilical cord blood was collected for the children at delivery. All the blood samples were processed by the Norwegian Institute of Public Health and DNA was extracted and stored following standard procedures.^4^

***Genotyping and quality control***

Blood samples in MoBa were genotyped over several research projects including HARVEST, SELECTIONpreDISPOSED, and NORMENT. About 238,001 samples were genotyped at 3 centers (Genomics Core Facility, Trondheim, Norway; deCODE Genetics, Reykjavik, Iceland; ERASMUS MC, Rotterdam, Netherlands) in 24 genotyping batches. Different selection criteria and genotyping arrays have been applied to those samples. To harmonize the complexity of the MoBa genotype data, QC was performed on the single nucleotide polymorphism (SNP) and individual level following the MoBaPsychGen pipeline,^5^ which includes pre-imputation QC, phasing, imputation, and post-imputation QC. Full details can be found here.^5^

***Maltreatment history***

Questions regarding parents’ own maltreatment history (physical, sexual, emotional, verbal abuse) during childhood and adulthood were asked during pregnancy (pregnancy week 15 and 30). When the children were 8 years old, mothers were also asked about their children’s physical abuse history (Supp. Table 1). All maltreatment variables were coded as binary (yes/no).

***Mental health and behavioral outcomes***

Parental anxiety and depression traits were measured using Hopkins Symptoms Checklist (SCL) during pregnancy week 15 and 30. Children’s mental health and neurodevelopment/behavior including traits of depression, anxiety, and attention deficit hyperactivity disorder (ADHD) were measured using Short Mood and Feelings Questionnaire (SMFQ),^6^ Screen for Child Anxiety Related Disorders (SCARED),^7^ and parent / teacher rating scale for disruptive behavior disorders (RS-DBD)^8^ respectively at age 8 years (Supp. Table 2). The children answered the same set of mental health and neurodevelopment/behavior questionnaires at age 14 years in addition to the Hopkins Symptoms Checklist used to assess depression and anxiety traits. However, for ADHD traits at age 14 years, mother-reported RS-DBD was used since the child-reported scale was not available. We created prorated scores for those with at least 80% item-level information by multiplying the raw scores with the total number of items in the mental health questionnaires and dividing by the number of items completed by the participants. The prorated scores were log transformed or binary coded if log transformation did not correct for skewness. The cut-offs that were used for parental SCL, mother-reported SCARED, and mother-reported SMFQ at age 8 years were mean score (prorated score / total items) ≥2, total score ≥2, and total score≥11 respectively.

***Statistical analyses***

We generated polygenic risk scores for each participant based on the GWAS of childhood maltreatment of 185,414 individuals^9^ using PLINK 1.9. After clumping (r2=0.01, 10000kb), 12 SNPs which were independently associated (at p=5 x 10^-8^) with childhood maltreatment in the GWAS were included and INFO score was > 0.95. All PRS generated were z-standardized.

Using log binomial and linear regressions, we examined associations of childhood maltreatment PRS in mothers (PRS_M_), fathers (PRS_F_), and children (PRS_C_) with their respective maltreatment experiences and mental health and behavioral outcomes including traits of anxiety, depression, and ADHD. To assess confounding by dynastic effects, we examined whether associations of PRS_C_ with mothers’ reports of their children’s physical abuse and child mental health and behavioral outcomes were attenuated with adjustment for parental genotype. All PRS models were adjusted for individual birth year, 20 PCs, and genotyping center and chip. The child PRS model was further adjusted for the child’s sex and clustering by Family ID using robust standard errors. To assess parental indirect effects, child mental health and behavioral outcomes were regressed on parental PRS adjusting for PRS_C_. The associations between PRS_C_ and mothers’ reports of their own childhood maltreatment history and mental health outcomes adjusting for PRS_M_ were used as negative control. Finally, we performed sensitivity analyses using PRS generated with the inclusion of all SNPs without using a p-value threshold for the PRS_C_ – child mental health and behavioral outcomes analyses adjusting for parental PRS. We also performed non-genetic regression models to examine associations of mothers’ reports of their children’s physical abuse at age 8 years and child mental health and behavioral outcomes at ages 8 and 14 years adjusting for relevant covariates. The covariates included birthweight, parity, multiple birth, mother’s smoking at age 8 years, mother reported father’s smoking at age 8 years, gestational age, maternal age at delivery, sex, mother reported father’s education at pregnancy week 15, mother’s education at age 8 years.

**Supplementary tables**

| **Role** | **Type of abuse** | **Timepoint** | **Questions** | **Options** |
| --- | --- | --- | --- | --- |
| Child | Physical | Y8 | Q6: In the course of the past 12 months, has your child been subjected to beating, kicking or other violence by adults? | Never / seldom / 2-3 times per month / once a week / many times per week |
| Mother | Physical | PW15 | Q139: Have you ever in your adult life been slapped, hit, kicked or bothered in any way physically? (you may cross off several) - during this pregnancy / last 6 months before pregnancy / earlier | Yes / no / don't remember |
| Mother | Physical | PW30 | Q129: Have you ever experienced any of the following? (Q3. You have been subjected to physical abuse) | No, never / yes, as a child (<18) / yes, as an adult (>=18) |
| Father | Physical | PW15 | Q73: Have you experienced any of the following during the last 12 months? (Q11: exposed to physical violence) | Yes / no |
| Mother | Sexual | PW15 | Q140: Have you ever been pressured or forced to have sexual intercourse? - during this pregnancy / last 6 months before pregnancy / earlier | No, never / yes, pressured / yes, forced with violence / yes, raped |
| Mother | Sexual | PW30 | Q129: Have you ever experienced any of the following? (Q4. You have been forced to have sexual intercourse) | No, never / yes, as a child (<18) / yes, as an adult (>=18) |
| Father | Sexual | PW15 | Q73: Have you experienced any of the following during the last 12 months? (Q10: forced into sexual activity) | Yes / no |
| Mother | Emotional | PW30 | Q129: Have you ever experienced any of the following? (Q1. Someone has over a long period of time systematically tried to subdue, degrade or humiliate you) | No, never / yes, as a child (<18) / yes, as an adult (>=18) |
| Mother | Verbal | PW30 | Q129: Have you ever experienced any of the following? (Q2. Someone has threatened to hurt you or someone close to you) | No, never / yes, as a child (<18) / yes, as an adult (>=18) |

***Supplementary Table 1*.** Maltreatment questions asked of the participants at various time points. Y8 is when the child was eight years of age. PW=pregnancy week. All variables were coded as binary for the final analyses.

***Supplementary Table 2.*** Validated psychological instruments used to measure mental health and behavioral outcomes. Y8 is when the child was eight years of age. Y14 is when the child was 14 years of age. PW=pregnancy week.

|  | **Child** | **Mother** | **Father** |
| --- | --- | --- | --- |
| Anxiety / Depression *(Hopkins Symptoms Checklist; SCL)* | Y14 (child report) | PW30 | PW15 |
| ADHD *(Parent / teacher rating scale for disruptive behavior disorders; RS-DBD)* | Y8 (mother report), Y14 (mother report) |  |  |
| Anxiety *(Screen for Child Anxiety Related Disorders; SCARED)* | Y8 (mother report), Y14 (child report) |  |  |
| Depression *(Short Mood and Feelings Questionnaire; SMFQ)* | Y8 (mother report), Y14 (child report) |  |  |

**Supplementary figures**

206,909 individuals with genetic data in the MoBa cohort that passed quality control

42,101 complete parent-offspring trios with consent available

42,101 unique children

10,742 siblings

738 (369 twin pairs)

3 (1 triplet)

36,618 unique mothers

36,612 unique fathers

***Supplementary Figure 1*** Cohort flowchart of study participants included in the final analytic sample.

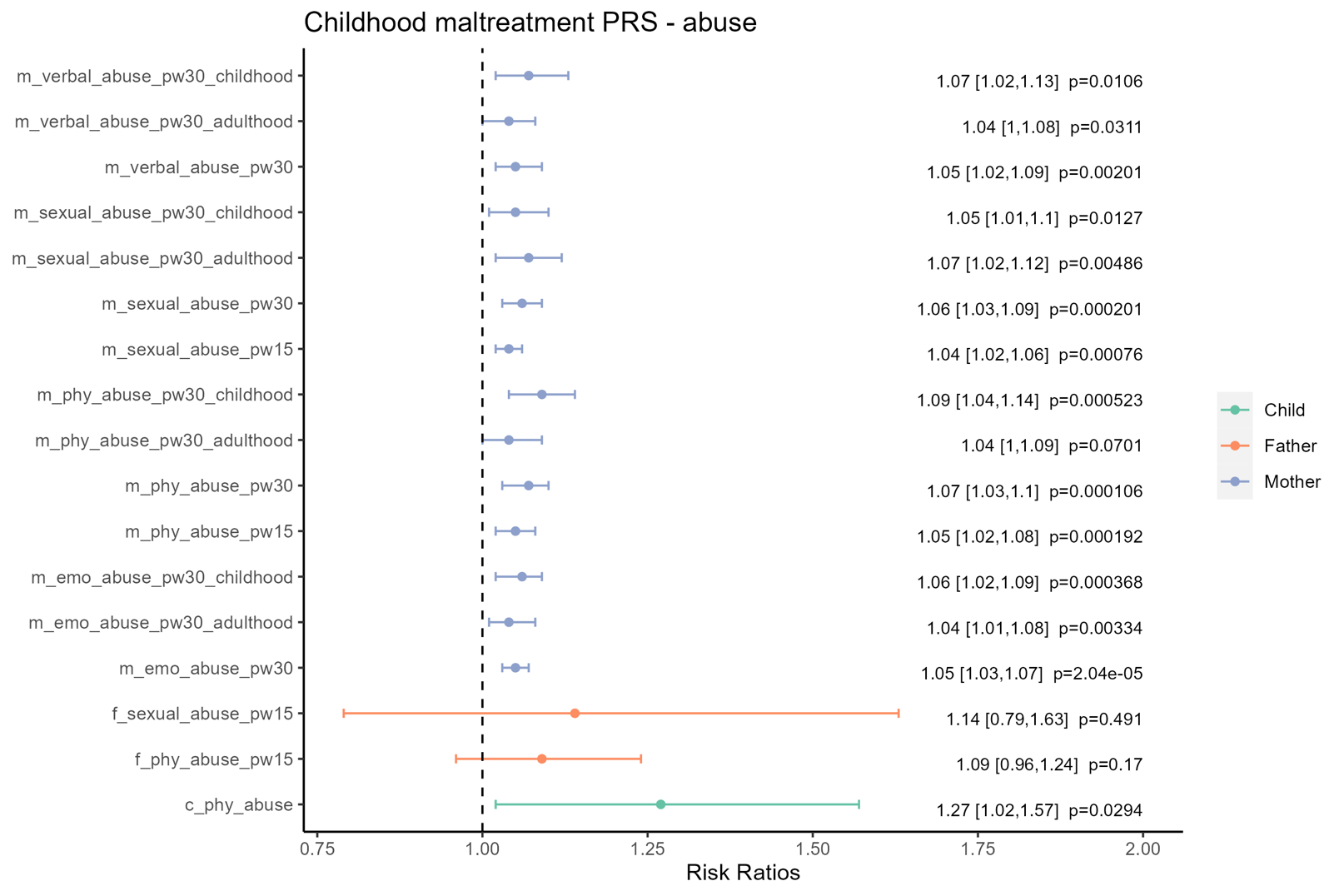

***Supplementary Figure 2*** Associations between PRS and maltreatment experiences in mothers, fathers, and children. Blue indicates associations between PRS_M_ and mothers’ reports of their own childhood and adulthood maltreatment experiences. Orange indicates associations between PRS_F_ and fathers’ reports of their own adult maltreatment experiences. Green indicates associations between PRS_C_ and mothers’ reports of their children’s maltreatment experiences. All PRS models were adjusted for individual birth year, 20PCs, genotyping center and chip. The child model was further adjusted for the child’s sex and accounted for family clustering using robust standard errors.

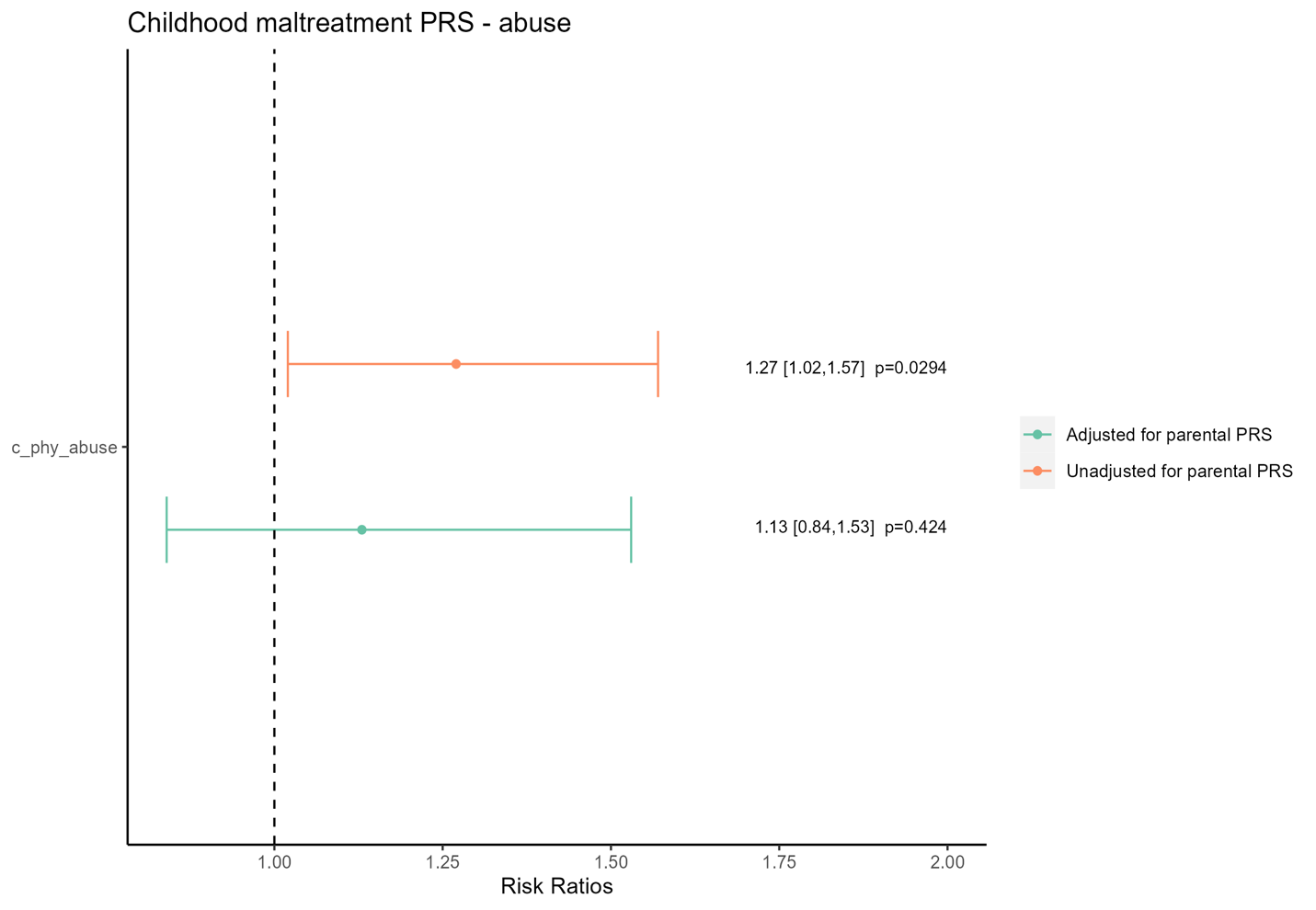

***Supplementary Figure 3*** Associations between PRS_C_ and mother’s report of child’s exposure to physical abuse measured at age 8 years before and after adjusting for parental PRS. Orange indicates associations before adjusting for parental PRS. Blue indicates associations after adjusting for parental PRS. All models were adjusted for individual birth year, 20PCs, genotyping center and chip, and the child’s sex. Clustering was accounted for using robust standard errors.

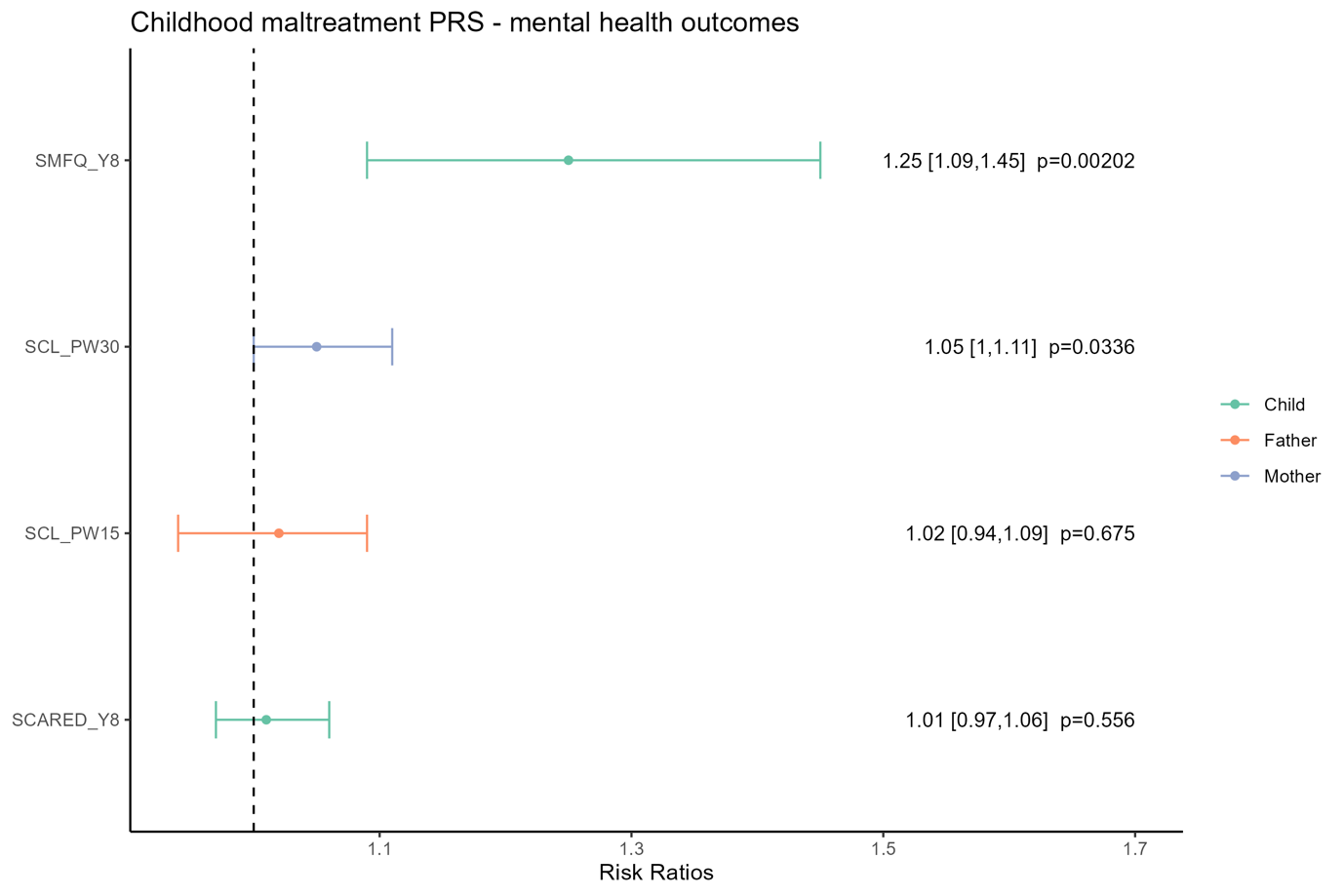

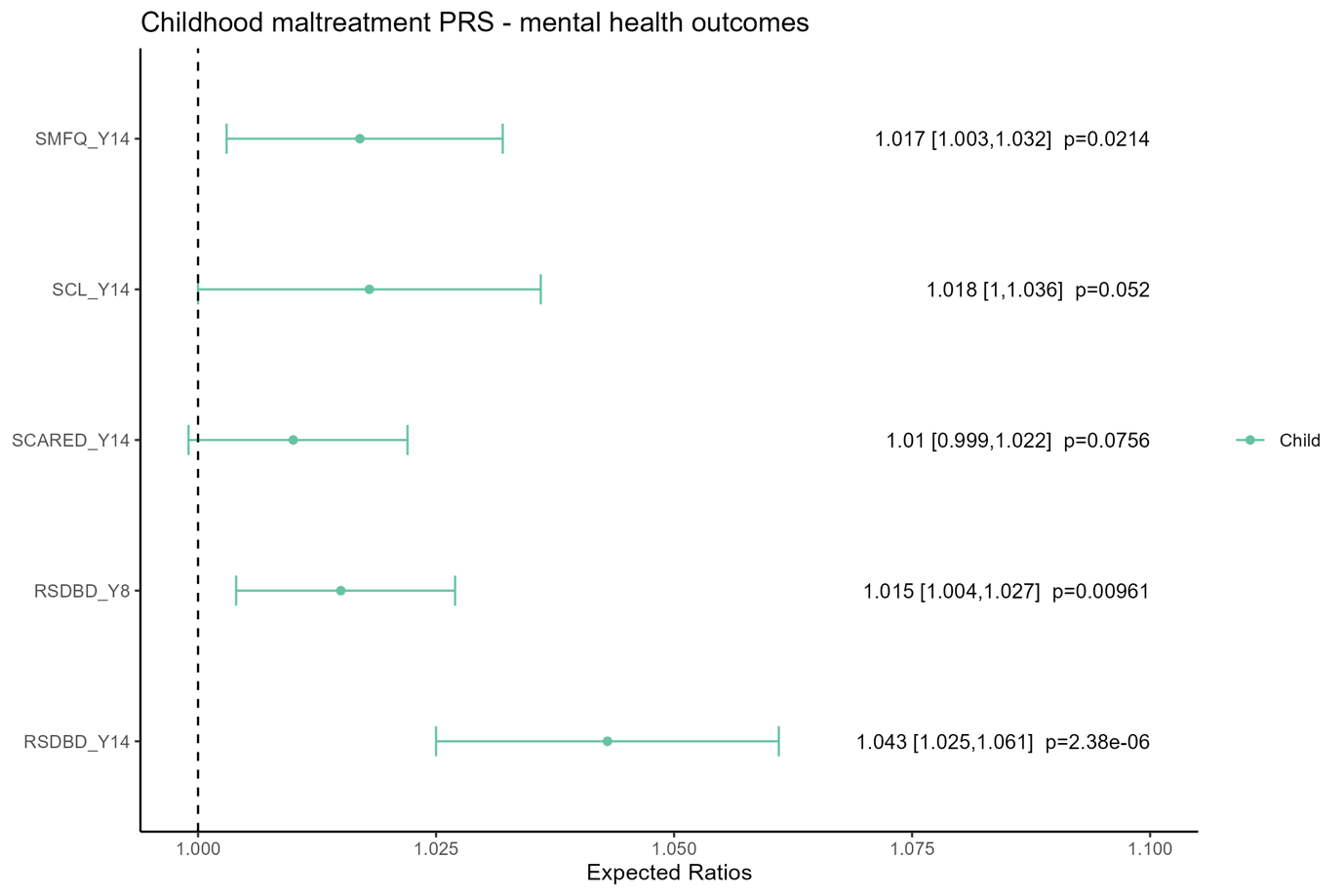
***Supplementary Figure 4*** Associations between PRS and mental health and behavioral outcomes in mothers, fathers, and children. **(a)** Log transformed outcomes. Effect estimates were reported as ratios of geometric means. **(b)** Binary outcomes. Blue indicates associations between PRS_M_ and mothers’ mental health outcomes during pregnancy. Orange indicates PRS_F_ and fathers’ mental health outcomes during pregnancy. Green indicates PRS_C_ and children’s mental health and behavioral outcomes at ages 8 and 14 years. All PRS models were adjusted for individual birth year, 20PCs, genotyping center and chip. The child model was further adjusted for the child’s sex and accounted for family clustering using robust standard errors. SCL= Hopkins Symptoms Checklist; SMFQ=Short Mood and Feelings Questionnaire; SCARED=Screen for Child Anxiety Related Disorders; RS-DBD=Parent / teacher rating scale for disruptive behavior disorders. Y8 is when the child was eight years of age. Y14 is when the child was 14 years of age. PW=pregnancy week.

4b.

4a.

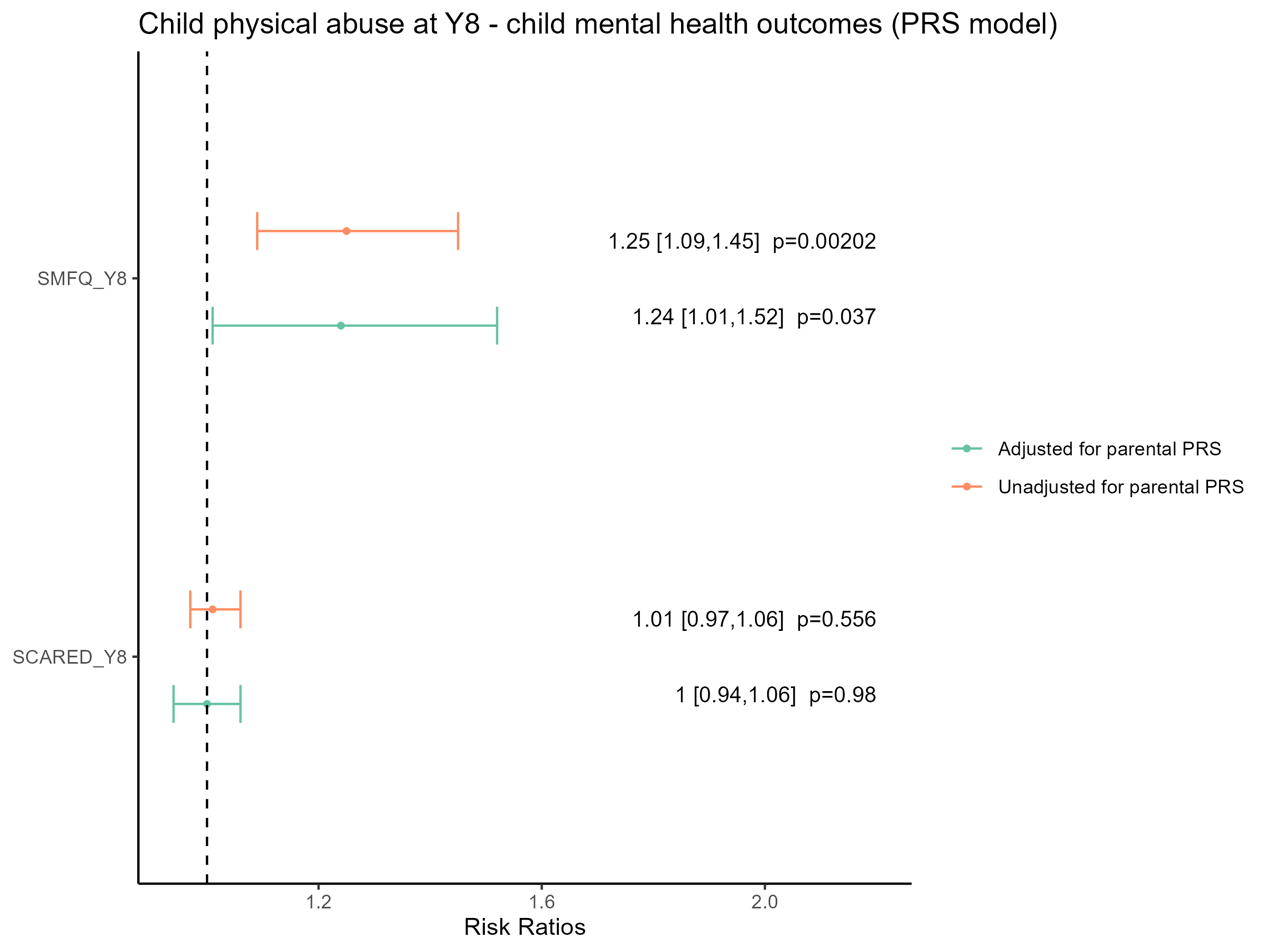

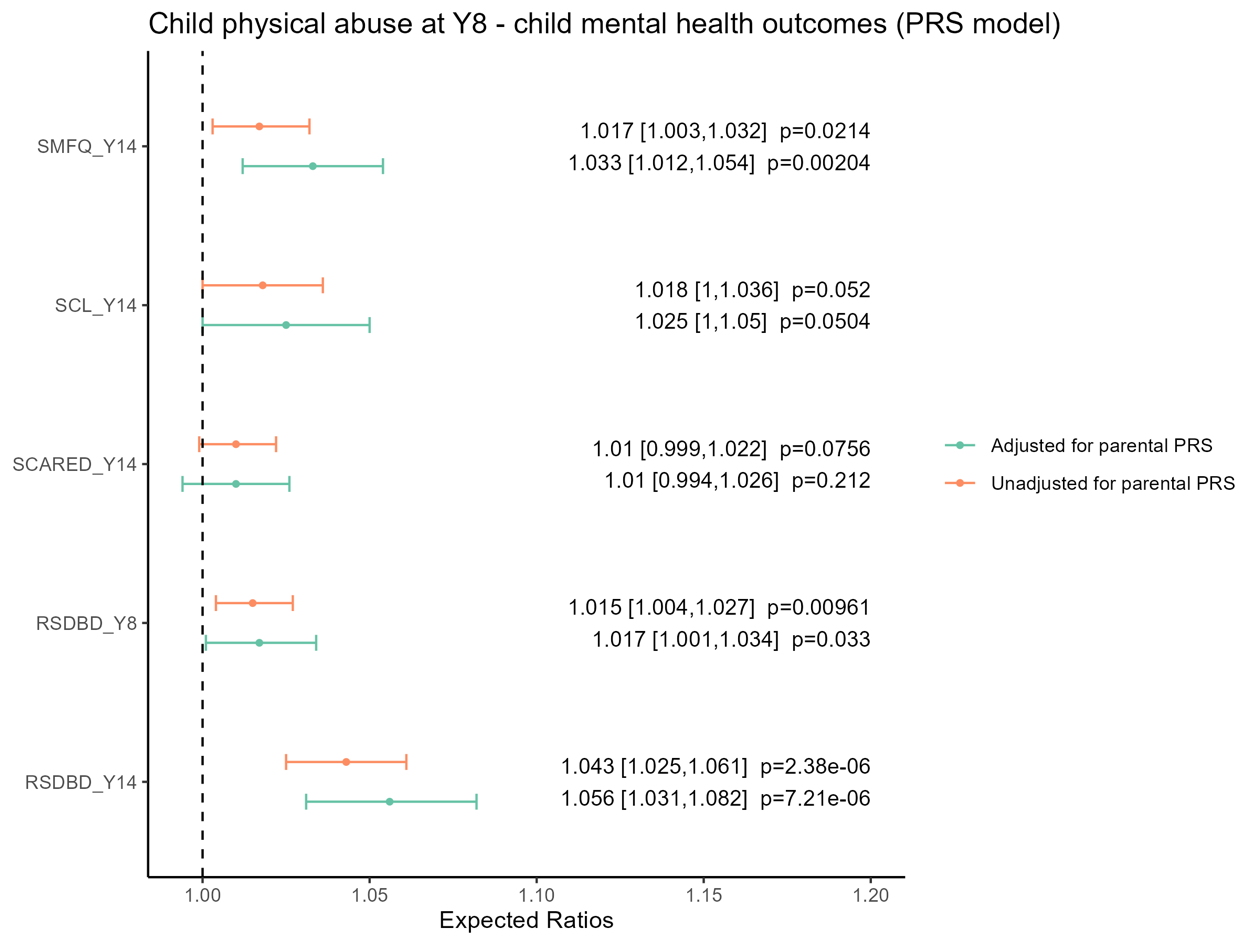
***Supplementary Figure 5*** Associations between PRS_C_ and children’s mental health and behavioral outcomes measured at ages 8 and 14 years before and after adjusting for parental PRS. **(a)** Log transformed outcomes. Effect estimates were reported as ratios of geometric means. **(b)** Binary outcomes. Orange indicates associations before adjusting for parental PRS. Green indicates associations after adjusting for parental PRS. All models were adjusted for individual birth year, 20PCs, genotyping center and chip, and the child’s sex. Clustering was accounted for using robust standard errors. SCL= Hopkins Symptoms Checklist; SMFQ=Short Mood and Feelings Questionnaire; SCARED=Screen for Child Anxiety Related Disorders; RS-DBD=Parent / teacher rating scale for disruptive behavior disorders. Y8 is when the child was eight years of age. Y14 is when the child was 14 years of age.

5b.

5a.

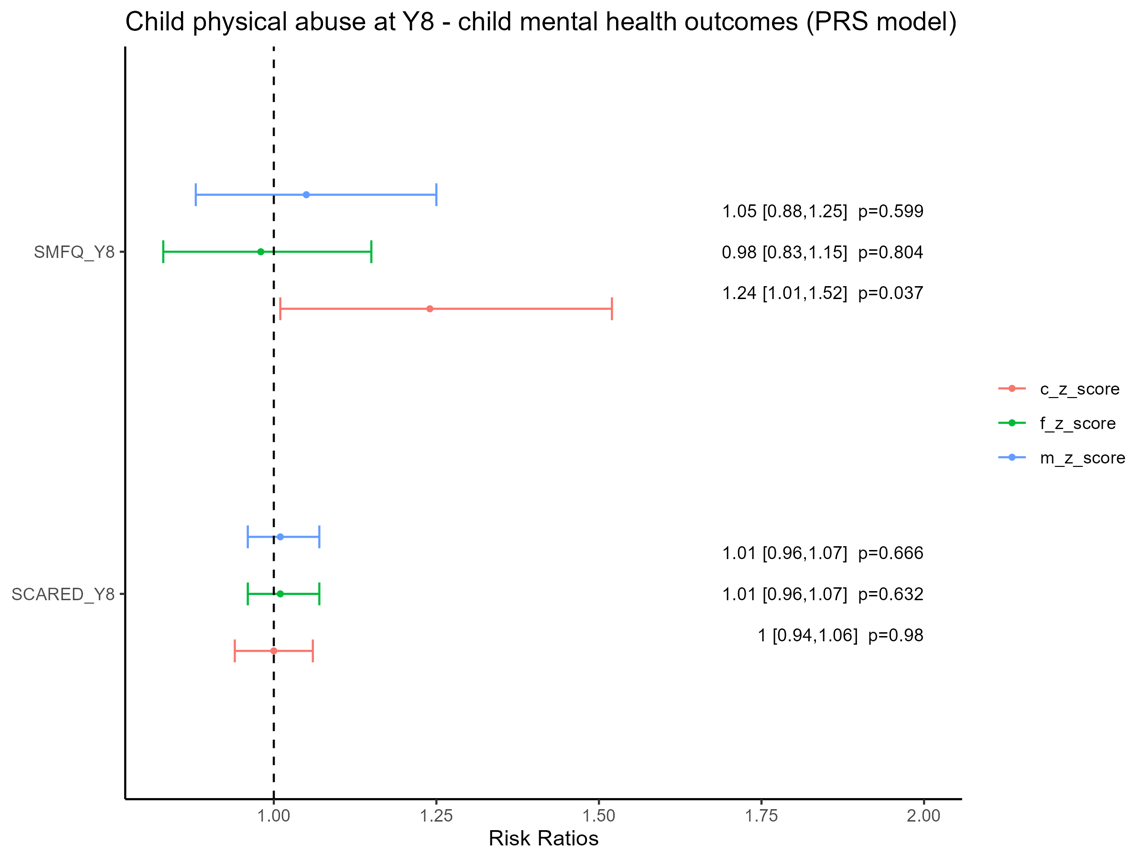

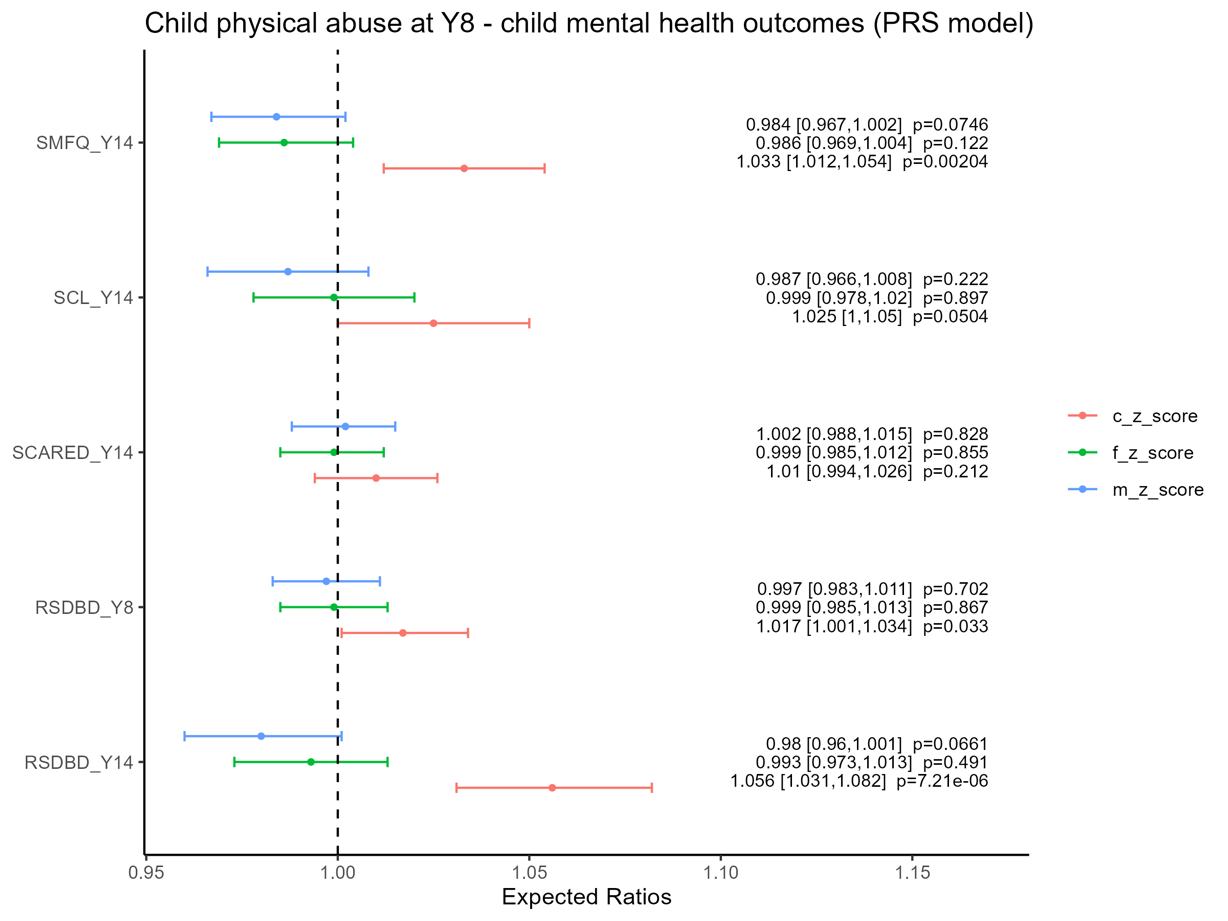
***Supplementary Figure 6*** Associations between PRS and children’s mental health and behavioral outcomes measured at ages 8 and 14 years after mutually adjusting for each other’s PRS. **(a)** Log transformed outcomes. Effect estimates were reported as ratios of geometric means. **(b)** Binary outcomes. Red indicates associations between PRS_C_ and children’s mental health and behavioral outcomes after adjusting for parental PRS. Green indicates associations between PRS_F_ and children’s mental health and behavioral outcomes after adjusting for PRS_M_ and PRS_C._ Blue indicates associations between PRS_M_ and children’s mental health and behavioral outcomes after adjusting for PRS_F_ and PRS_C_. Models were adjusted for individual birth year, 20PCs, genotyping center and chip. The child model was further adjusted for the child’s sex and accounted for family clustering using robust standard errors. SCL= Hopkins Symptoms Checklist; SMFQ=Short Mood and Feelings Questionnaire; SCARED=Screen for Child Anxiety Related Disorders; RS-DBD=Parent / teacher rating scale for disruptive behavior disorders. Y8 is when the child was eight years of age. Y14 is when the child was 14 years of age.

6b.

6a.

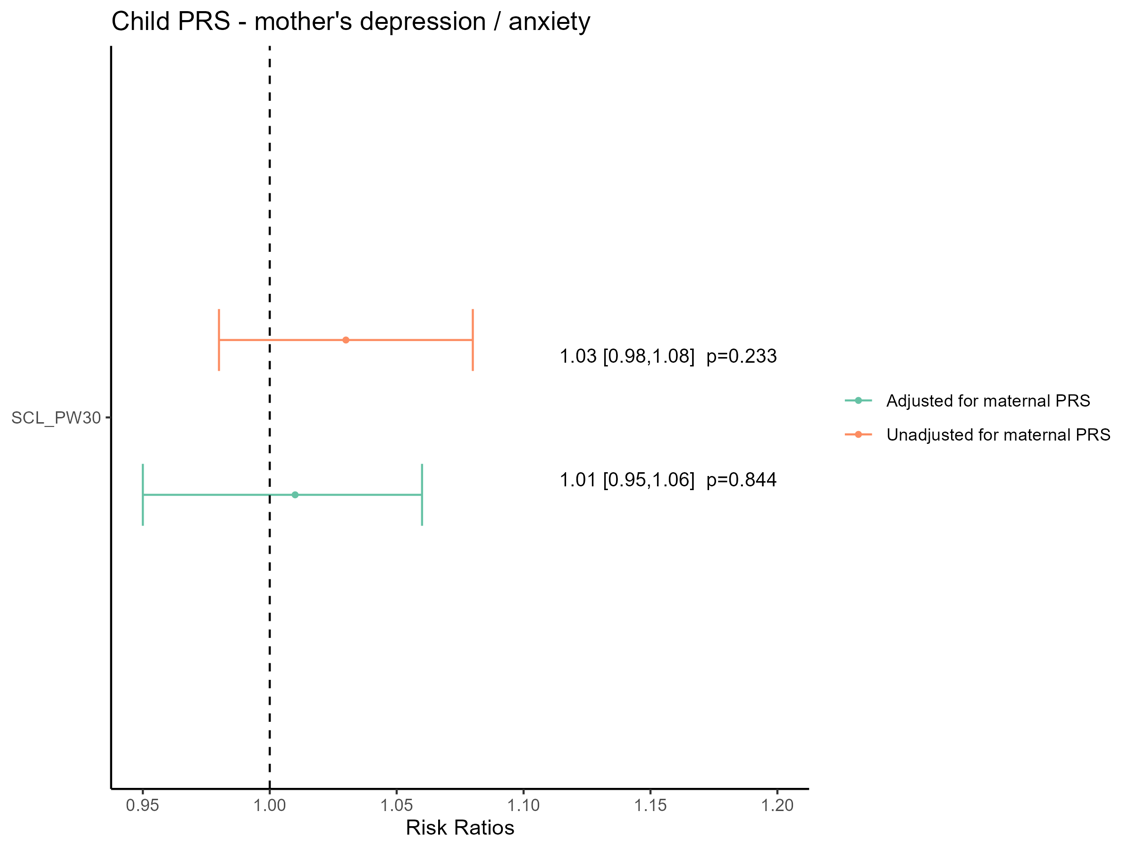

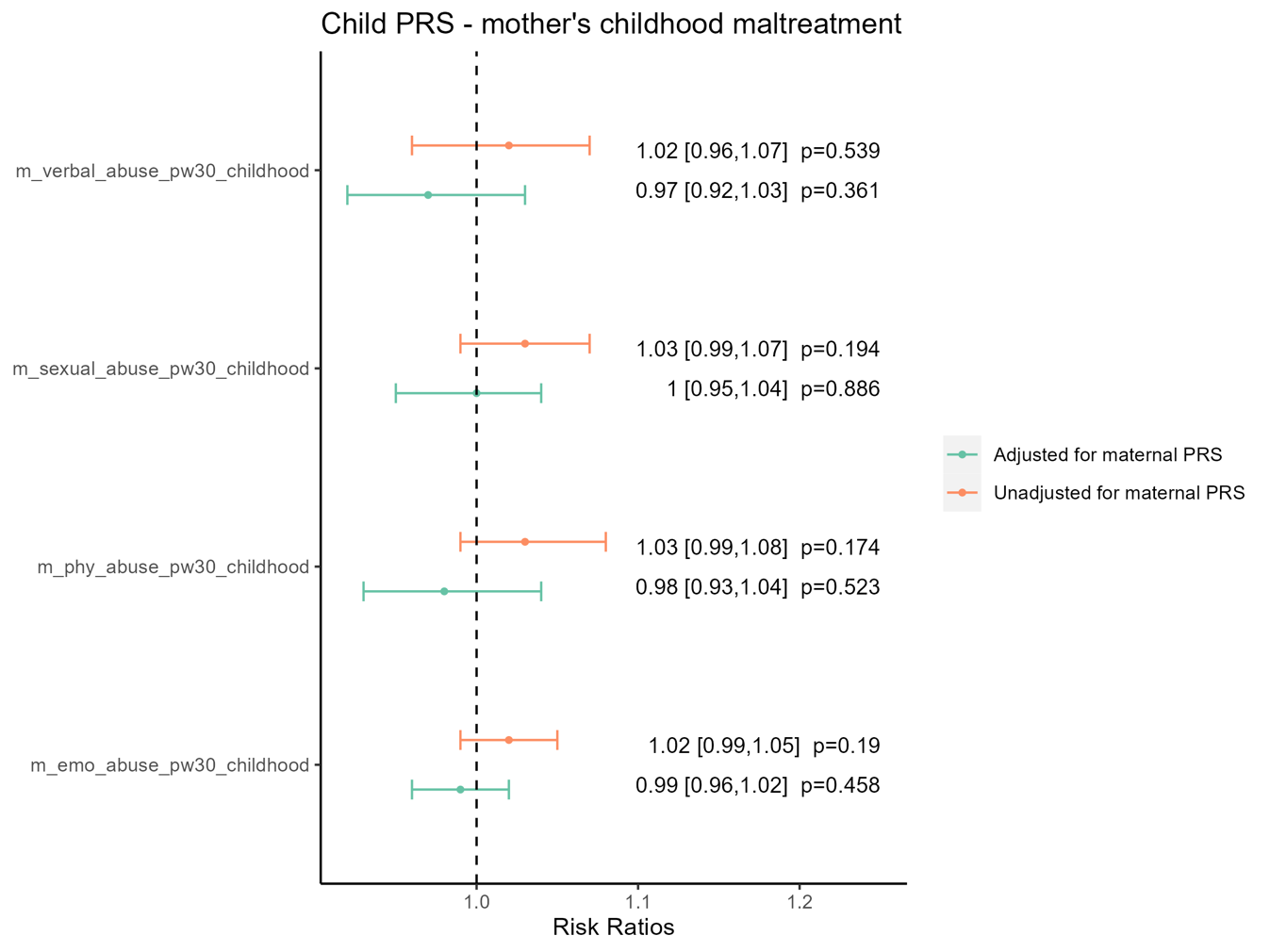
***Supplementary Figure 7*** Associations between PRS_C_ and mothers’ childhood maltreatment history and mental health outcomes measured during pregnancy before and after adjusting for maternal PRS. **(a)** Maternal childhood maltreatment history. **(b)** Maternal mental health outcomes during pregnancy. Orange indicates associations before adjusting for maternal PRS. Green indicates associations after adjusting for maternal PRS. All models were adjusted for individual birth year, 20PCs, genotyping center and chip, and the child’s sex. Clustering was accounted for using robust standard errors. SCL= Hopkins Symptoms Checklist; PW= pregnancy week.

7b.

7a.

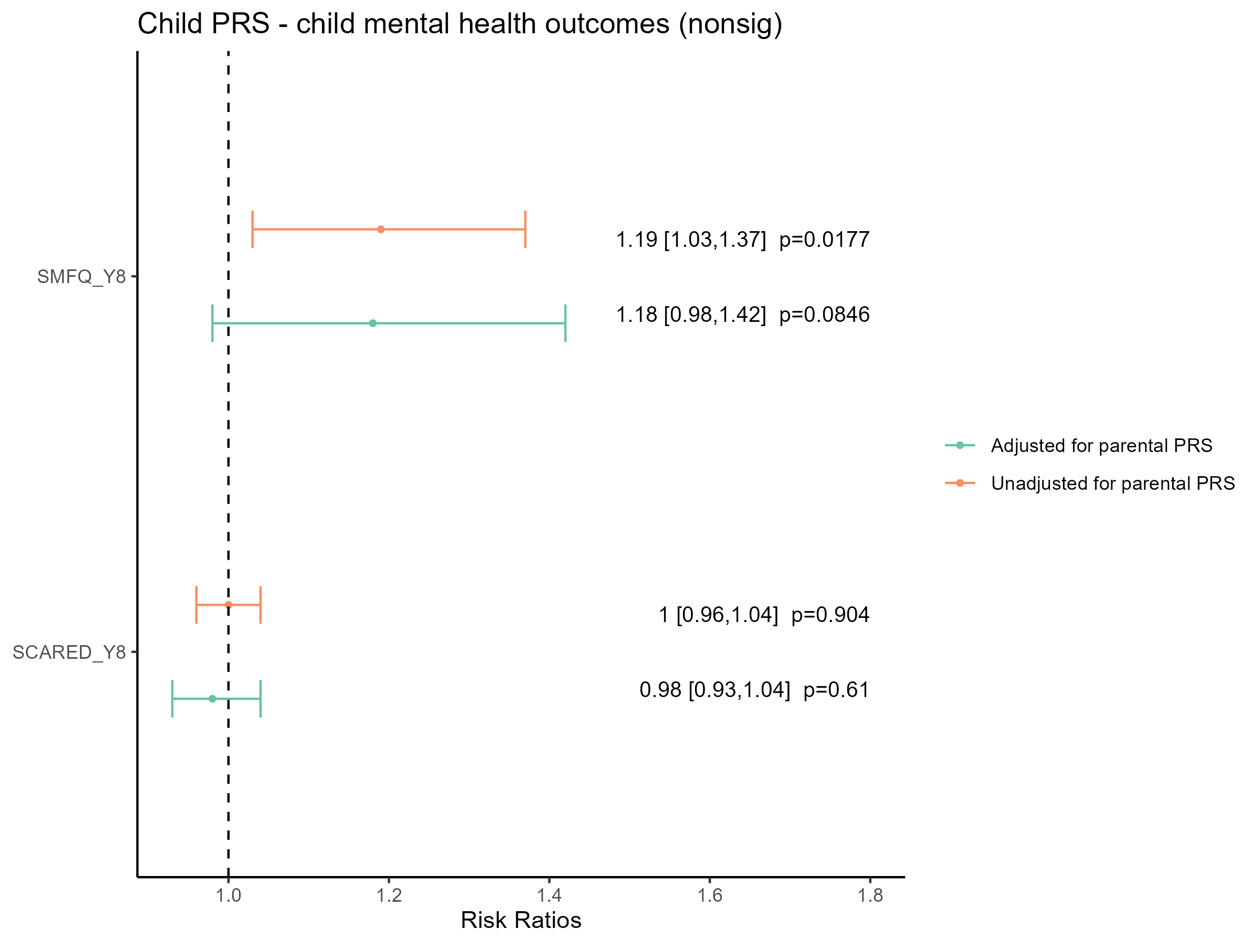

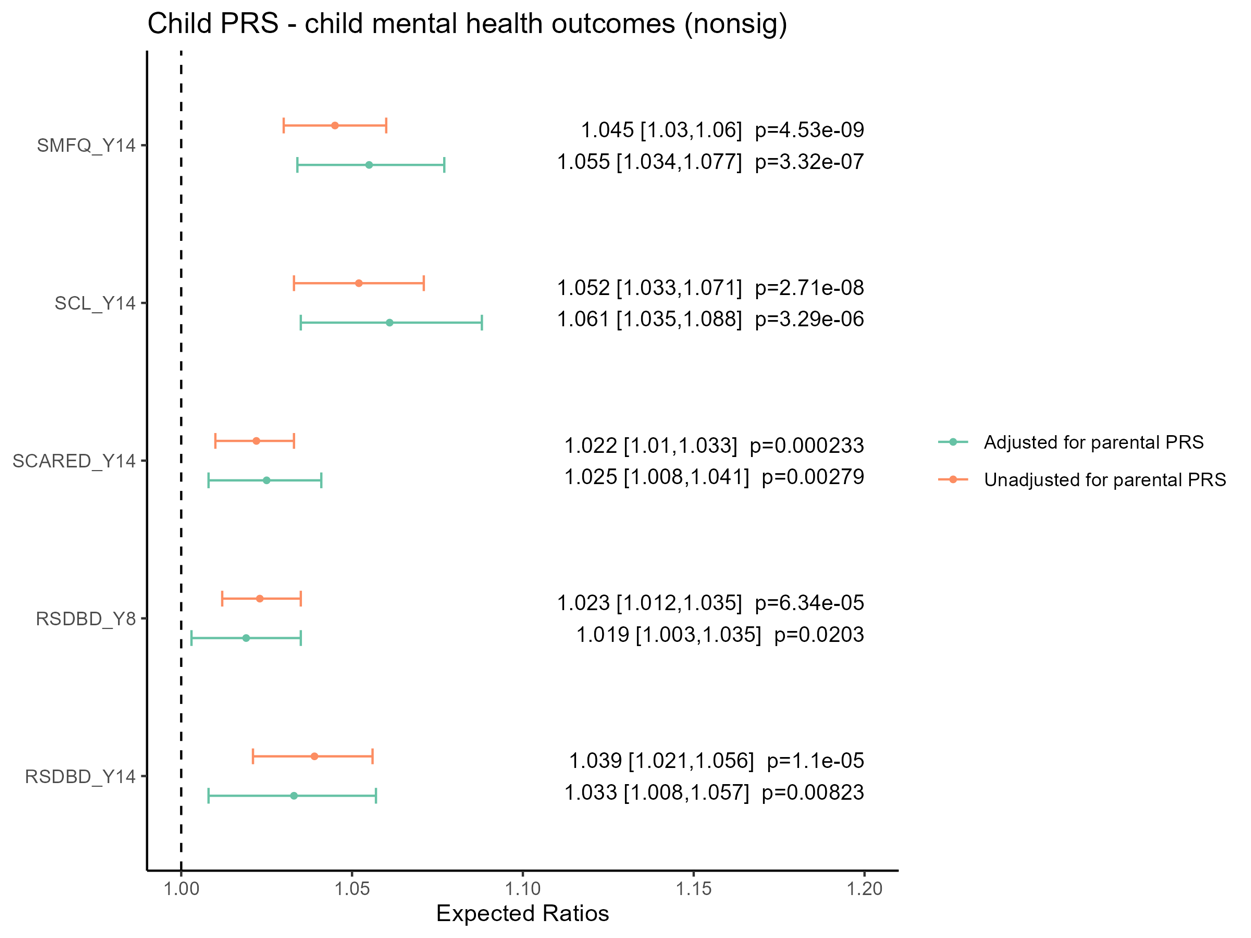
***Supplementary Figure 8*** Associations between PRS_C_ and children’s mental health and behavioral outcomes measured at ages 8 and 14 years before and after adjusting for parental PRS using PRS generated with the inclusion of all SNPs without specifying a p-value threshold. **(a)** Log transformed outcomes. Effect estimates were reported as ratios of geometric means. **(b)** Binary outcomes. Orange indicates associations before adjusting for parental PRS. Green indicates associations after adjusting for parental PRS. All models were adjusted for individual birth year, 20PCs, genotyping center and chip, and the child’s sex. Clustering was accounted for using robust standard errors. SCL= Hopkins Symptoms Checklist; SMFQ=Short Mood and Feelings Questionnaire; SCARED=Screen for Child Anxiety Related Disorders; RS-DBD=Parent / teacher rating scale for disruptive behavior disorders. Y8 is when the child was eight years of age. Y14 is when the child was 14 years of age.

8b.

8a.

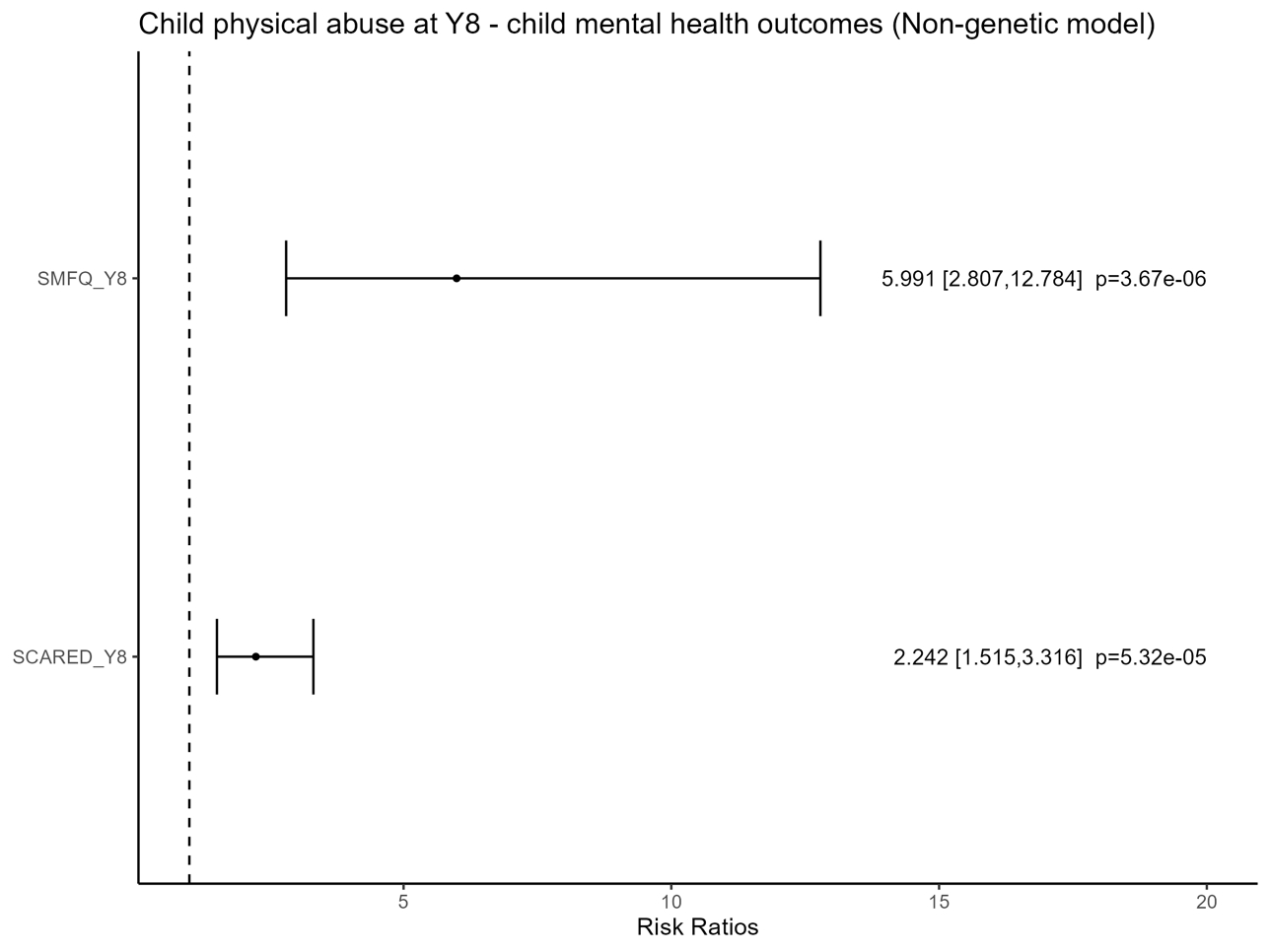

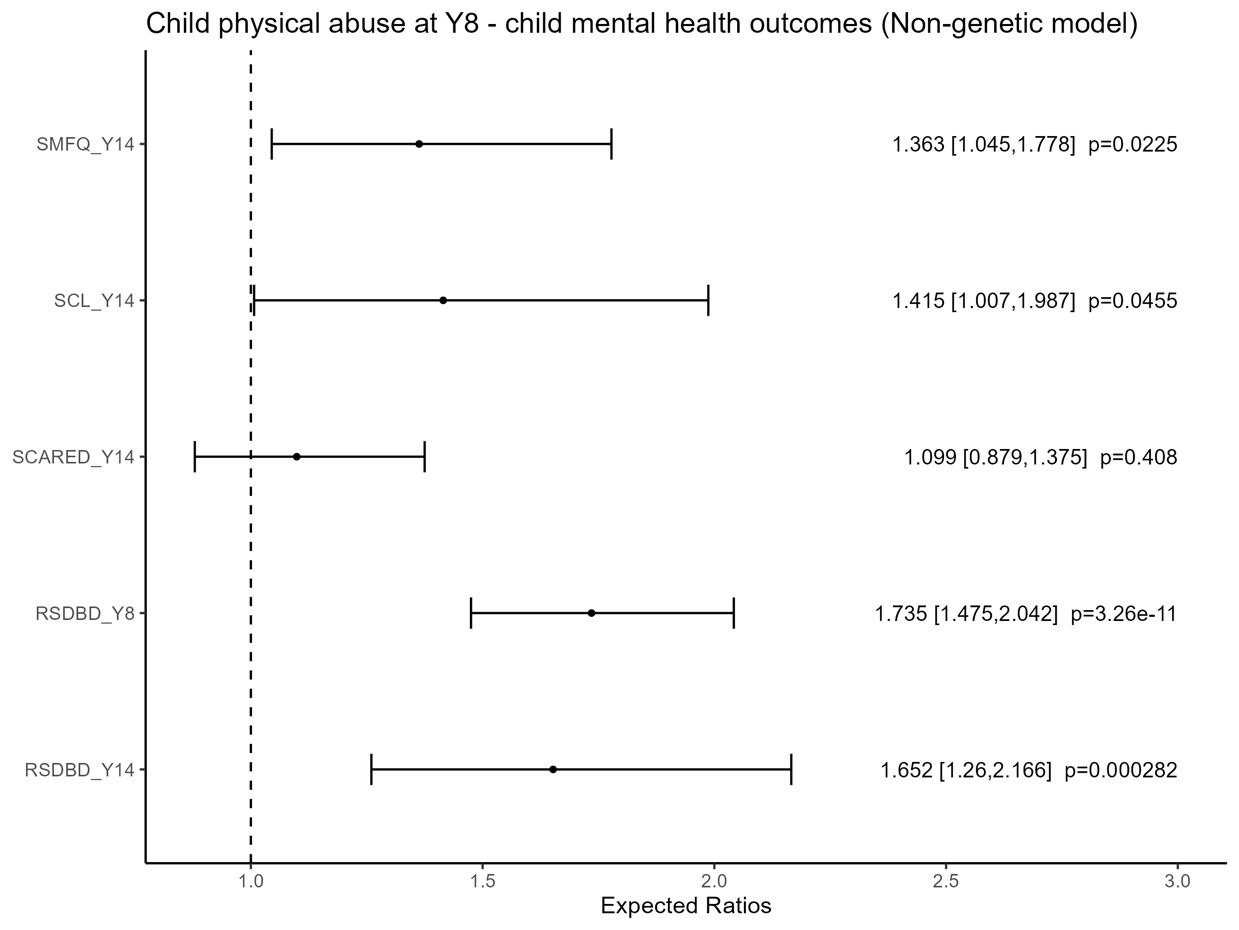
***Supplementary Figure 9*** Associations between mothers’ reports of children’s physical abuse at age 8 years and children’s mental health and behavioral outcomes measured at ages 8 and 14 years. **(a)** Log transformed outcomes. Effect estimates were reported as ratios of geometric means. **(b)** Binary outcomes. Models were adjusted for birthweight, parity, multiple birth, mother’s smoking at Y8, mother reported father’s smoking at Y8, gestational age, maternal age at delivery, the child’s sex, mother reported father’s education at PW15, mother’s education at Y8. Clustering was accounted for using robust standard errors. SCL= Hopkins Symptoms Checklist; SMFQ=Short Mood and Feelings Questionnaire; SCARED=Screen for Child Anxiety Related Disorders; RS-DBD=Parent / teacher rating scale for disruptive behavior disorders. Y8 is when the child was eight years of age. Y14 is when the child was 14 years of age.

9b.

9a.
